# Adaptor protein 2 sigma subunit (*AP2S1*) variants associated with neurodevelopmental disorders

**DOI:** 10.1101/2024.07.22.24310683

**Authors:** Mark Stevenson, Asha L. Bayliss, Victoria J. Stokes, Katherine A. English, Kreepa G. Kooblall, Roman Fischer, Raphael Heilig, Iolanda Vendrell, Maria E. W. A. Albers, Meghan Bartos, Amber Begtrup, Alexia Bourgois, Rebecca Buchert, David J. Carey, Deanna A. Carere, Amanda Carnevale, Kristl G. Claeys, Benjamin Cogne, Gregory Costain, Nicole de Leeuw, Anne-Sophie Denommé-Pichon, Elizabeth J. Donner, Eftychia Drogouti, David A. Dyment, Balram Gangaram, Tobias B. Haack, Jeremy S. Haley, Solveig Heide, Ralf A. Husain, Bertrand Isidor, Louise Izatt, Adeline Jacquinet, Jane Juusola, Juliette J. Kahle, Boris Keren, Eric W. Klee, Evgenia Kokosali, Brendan C. Lanpher, Erica L. Macke, Elysa J. Marco, Kirsty McWalter, Bryce A. Mendelsohn, Aubrey Milunsky, Matthew Osmond, Amelie Piton, Angelika Riess, Valentin Ruault, Patrick Rump, Sarah Schuhmann, Amelle L. Shillington, Diane T. Smelser, Lot Snijders Blok, Frederic Tran Mau-Them, Christos Tsakalidis, Abigail Turnwald, Koen L. I. Van Gassen, Kristof Van Schil, Georgia Vasileiou, Marissa Vawter-Lee, Marjolaine Willems, Marjolein H. Willemsen, Lily C. Wong-Kisiel, Antje Wonneberger, Ioannis Zaganas, Genomics England Research Consortium, Fadil M. Hannan, Kate E. Lines, Rajesh V. Thakker

## Abstract

*Adaptor-Related Protein Complex 2 Sigma-1 Subunit* (*AP2S1*) encodes AP2σ2, which forms part of the heterotetrameric AP2 complex that is composed of α, β2, μ2, and σ2 subunits and has a pivotal role in clathrin-mediated endocytosis (CME)^1–3^. *AP2S1* variants involving the Arg15 residue are associated with familial hypocalciuric hypercalcaemia type 3 (FHH3)^1,4–6^. Here, we report 5 different *AP2S1* variants (AP2σ2: p.Arg10Trp, p.Arg10Gln, p.Lys18Glu, p.Lys18Asn and p.Arg61His) in 26 patients with neurodevelopmental delay, of whom >70% had epilepsy, 50% had brain abnormalities, and none had hypercalcaemia. All 5 variants decreased cell viability, 4 reduced CME transferrin uptake, and 4 disrupted interactions with other AP2 complex subunits, thereby affecting AP2 formation. Furthermore, AP2σ2 p.Arg10Trp had reduced interactions with 44 human proteins including intersectin 1, a component required for clathrin-coated pit formation and synaptic vesicle dynamics in neurones. Thus, our results show that AP2σ2 variants may disrupt CME and be associated with neurodevelopmental disorders.

## Main Text

We identified 26 individuals (age range: 0-55 years) who had neurodevelopmental disorders (NDD) and were heterozygous for AP2σ2 variants p.Arg10Trp (R10W), p.Arg10Gln (R10Q), p.Lys18Glu (K18E), p.Lys18Asn (K18N) or p.Arg61His (R61H) (Table 1 and Supplementary Fig. 1). All variants occurred *de novo* in unrelated patients with similar phenotypes, which comprised: developmental delay (n=26); epilepsy (n=19); delayed speech (n=20); atypical behaviour including autism (n=8); attention deficit hyperactivity disorder (ADHD; n=7); abnormal gait and coordination (n=8); hypotonia (n=12); and brain abnormalities that included decreased cerebral volume and/or corpus callosum hypoplasia (n=10 of 20 investigated cases) (Table 1). The NDD patients (21 of 26 investigated cases) did not have hypercalcaemia which is the hallmark of FHH3^1^. These AP2σ2 variants were not found in the gnomAD database (v.4.1.0) of >800,000 samples^7^ with the exception of R10Q and R61H which occurred in 7/1,461,884 and 1/1,461,870 alleles, respectively. Moreover, these rare variants arose in residues R10, K18 and R61 that are evolutionarily conserved in AP2σ2 orthologues, and to varying degrees in AP1, AP3 and AP4 sigma subunit paralogues (Supplementary Fig. 2), thereby suggesting they have critical roles in function, and variant pathogenicity prediction tools indicated that these variants were likely deleterious (e.g., CADD scores >25) (Supplementary Table 1).

**Table 1.**
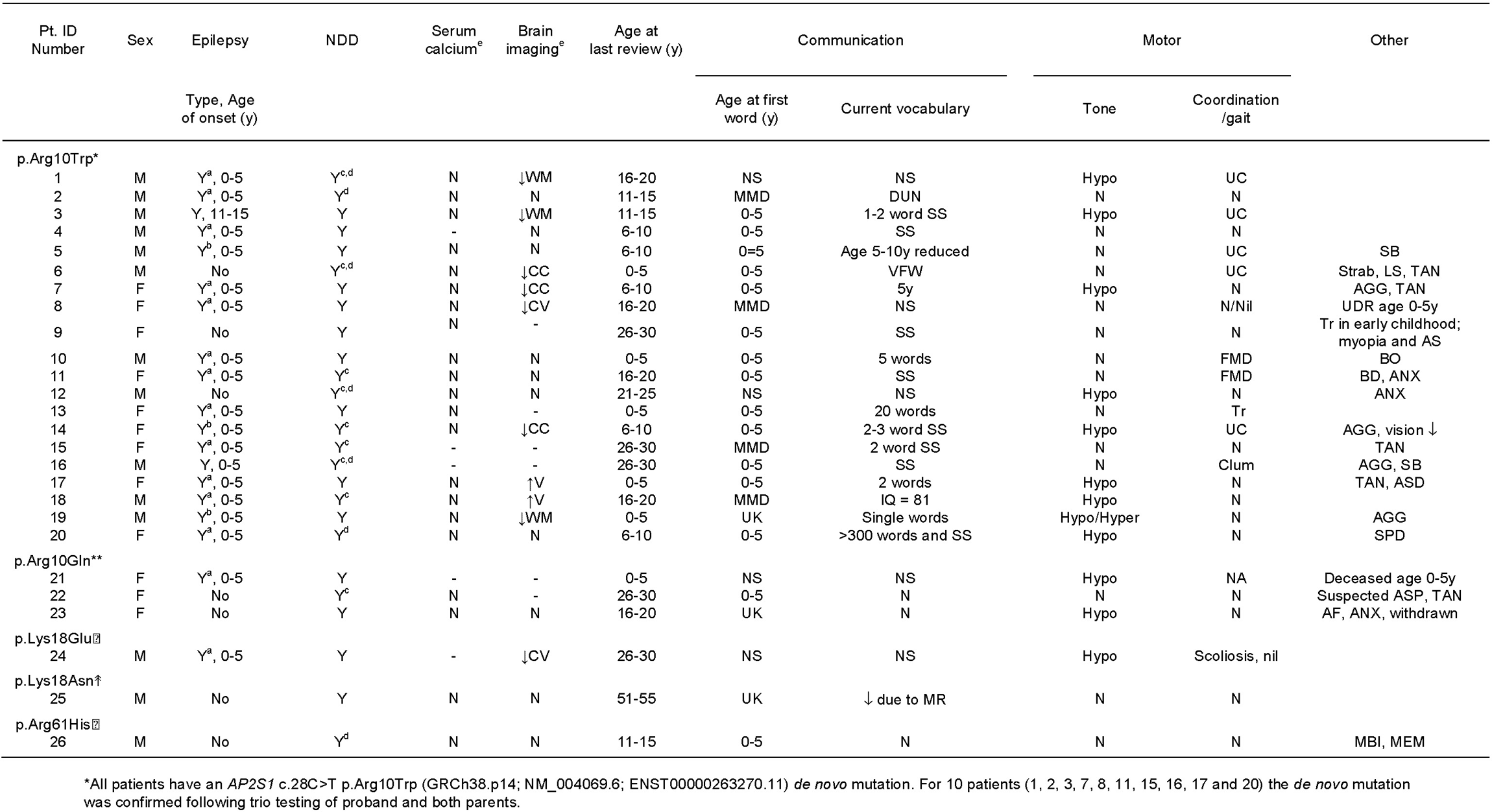
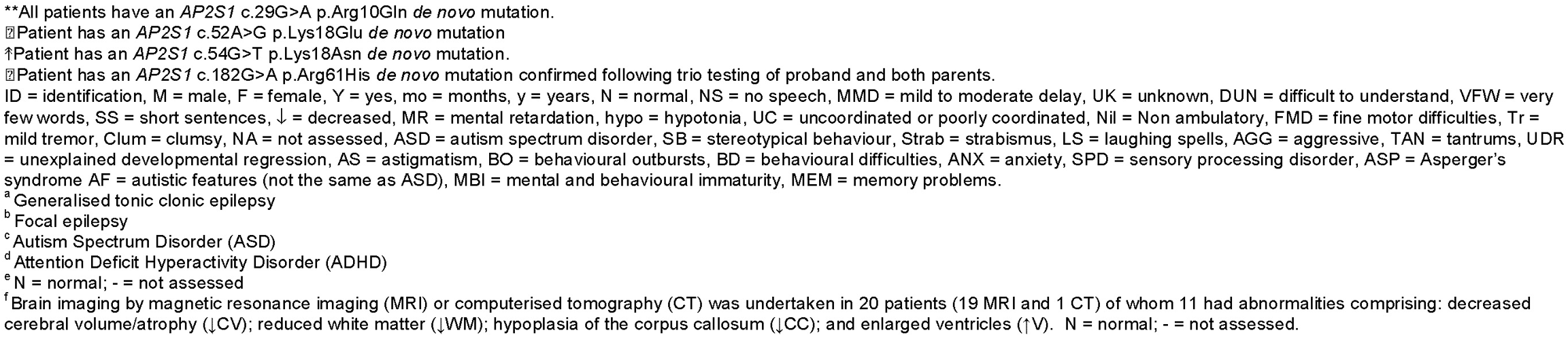
Clinical features of patients with neurodevelopmental delay (NDD)

To investigate deleterious consequences of these NDD-associated *AP2S1* variants, we initially assessed their effects on cell viability and CME, as these have been previously reported to be decreased by FHH3-associated AP2σ2-R15 variants^1,4,8^. Use of HeLa cells stably expressing HA-tagged wild-type (WT) (HeLa-AP2σ2H-WT) or variant (HeLa-AP2σ2H variant (-R10W, -R10Q, -R15L, -K18E, -K18N and -R61H)) *AP2S1* constructs (Supplementary Fig. 3), revealed that all 5 NDD-AP2σ2 variants and the FHH3-R15L variant were associated with reduced cell viability, compared to WT (Fig. 1a and Supplementary Fig. 4), thereby suggesting that these AP2σ2 variants may be pathogenic. We therefore assessed the effects of these NDD-associated AP2σ2 variants on CME, in which AP2 plays a central role in linking membrane associated cargo proteins^9^ with a dileucine-based or tyrosine-based motif (Fig. 1b and 1c) to clathrin and forming clathrin coated vesicles. The sites that bind dileucine-based and tyrosine-based endocytic cargo motifs are located on AP2σ2 and AP2μ2, respectively, and become available only in the unlatched and open conformations (Fig. 1b)^3,10,11^. This suggested that the NDD-associated AP2σ2 variants could be altering CME by disrupting binding to the dileucine-based motif, in a similar manner to that reported for the FHH3-associated variants, R15L, R15H and R15C, which decrease CME of the CaSR that has a dileucine endocytic motif^1,8^. In support of this both the R10 and R61 residues, which are involved in NDD-associated variants, were found in close proximity to the R15 residue in the AP2 open conformation (Fig. 1c), with R10 binding to the methionine (M) of the dileucine-based cargo protein in the unlatched conformation. However, investigation of the effects of the NDD-associated AP2σ2-R10W variant on CaSR signalling in HEK293 cells stably expressing the FLAG-tagged CaSR (HEK293-CF) and transfected with HA-tagged AP2σ2 (-WT or -R10W), revealed that the R10W variant did not affect CaSR signalling (Supplementary Fig. 5a and 5b), which is similar to our previous report showing that the AP2σ2-R61H variant does not alter CaSR signalling^12^, and consistent with normal plasma calcium concentrations in these patients with NDD (Table 1). These findings indicated that NDD-associated AP2σ2 variants were unlikely to alter CME by disruptions of binding to the dileucine-based motif, and we therefore investigated their effects on uptake of transferrin (Tf) by the transferrin receptor (TfR), which is a validated cargo protein that has a tyrosine-based motif for CME^13^, using HeLa cells.

**Figure 1.**
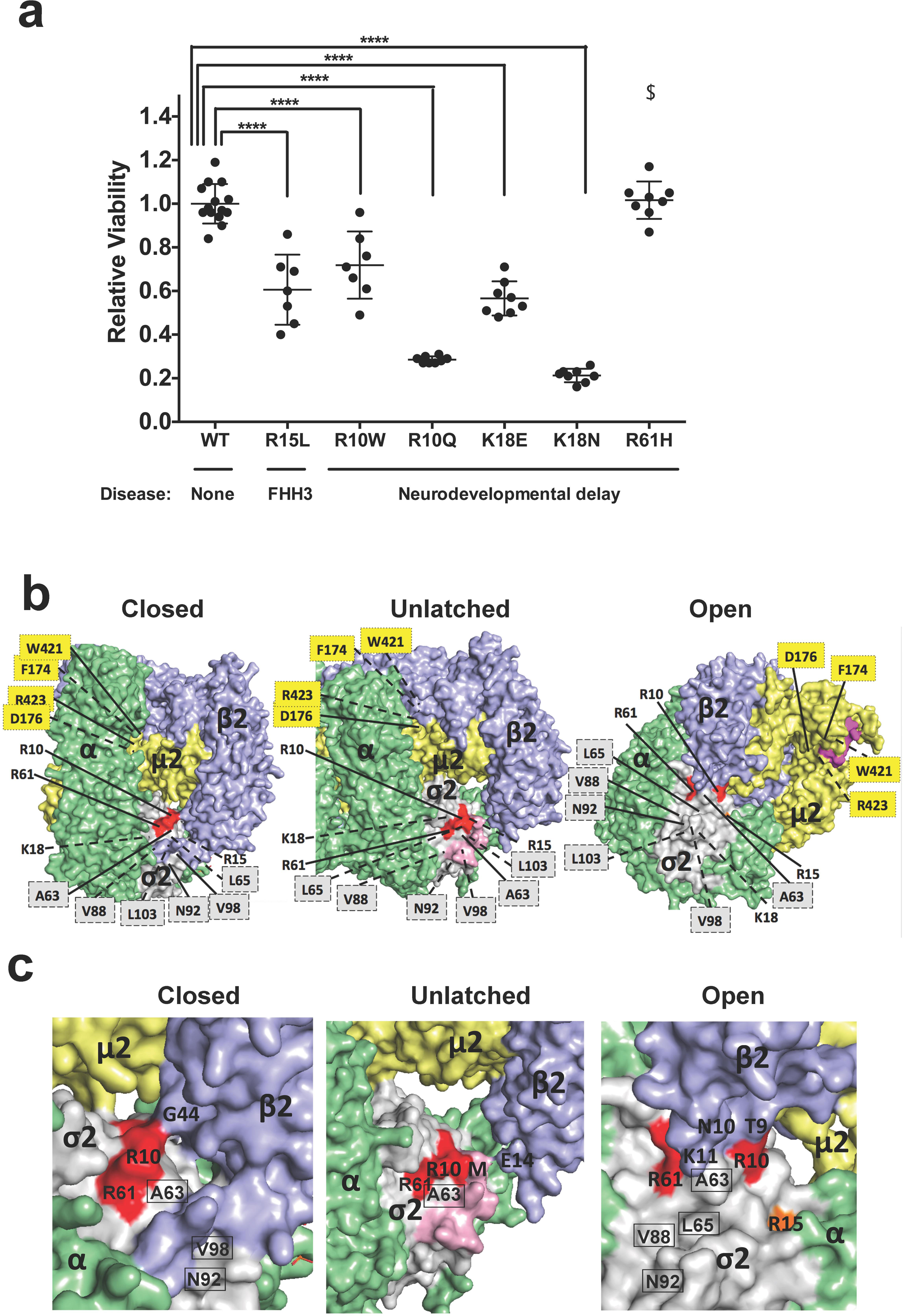
Viability of cells stably expressing AP2σ2-WT and -variant constructs, and locations of AP2σ2 residues associated with NDD- and FHH3-variants in the AP2 complex. **a,** Relative cell viability at 72h of HeLa cell lines stably expressing AP2σ2-WT (HeLa-AP2σ2H-WT), and AP2σ2-variants (HeLa-AP2σ2H -R10W, -R10Q, -R15L, -K18E, -K18N and -R61H). Viability rates were normalized such that WT cells reached 1.0 on day 3. Cells stably expressing the AP2σ2 variants R15L, R10W, R10Q, K18E and K18N had significantly reduced cell viability compared to WT by day 3, and those expressing R61H ($) showed significantly reduced viability at day 5 (Supplementary Figure 4). Error bars represent SD. Data was analysed using ANOVA and Tukey’s multiple comparisons test. ****p<0.0001. **b,** Locations of AP2σ2 residues whose variants are associated with NDD (R10, K18 and R61) and FHH3 (R15) are shown in red and orange respectively, on surface representations of AP2 complex with AP2α (green), AP2β2 (blue), AP2σ2 (grey), and AP2μ2 (yellow) subunits in the closed, unlatched and open conformations reported by crystallography studies^3,10,11^. The cargo protein RM(phosphoS)QIKRLLSE peptide from CD4 representing the dileucine ([ED]xxxL[LI]) endocytic cargo motif is shown in pink, and the DYQRLN peptide from TGN38 representing the tyrosine (YxxΦ) endocytic cargo motif is shown in magenta, in the unlatched and open conformations, respectively. The cargo dileucine-motif binds to a hydrophobic pocket formed by AP2σ2 residues that include A63, L65, V88, N92, V98 and L103 (grey boxes)^10^; whilst the cargo tyrosine-motif interacts with the AP2μ2 residues F174, D176, W421 and R423 (yellow boxes)^11,35^. In the closed conformation, AP2α and AP2β2 form the bulk of the exterior surface by surrounding the AP2σ2 and AP2μ2 subunits, which are blocked from interactions with the dileucine- and tyrosine-based motifs of cargo proteins. In the unlatched conformation, the N-terminus of AP2β2 is displaced, thereby allowing the dileucine-based motif to bind, while the tyrosine-based motif binding site remains blocked. In the open conformation, the C-terminus of the AP2μ2 subunit undergoes changes in orientation to allow the tyrosine-based motif to bind^3,10,11^. TfR uptake is largely dependent on Ap2μ2 which binds to a tyrosine based motif in the TfR cytoplasmic tail^36,37^, and following phosphorylation of Ap2μ2-Thr156 induces a conformation change that reveals the binding site with uptake into coated pits^38^. The R15 residue is buried within the subunit in the closed and unlatched conformations, and the K18 residue is buried in all 3 conformations, and therefore they are not visible from the surface. Residues on the surface are indicated with a solid black line, while residues that are not visible on the surface are indicated with a dotted black line. **c**. Zoomed in views showing locations of the AP2σ2 R10 and R61 residues, and the R15 residue in relation to the dileucine motif of the cargo peptide and the AP2β2 subunit. The AP2σ2 R10 is in contact with AP2β2 G44 in the closed conformation, but in the unlatched conformation it is in contact with the cargo methionine (M) residue which is in close contact with AP2β2 E14, and in the open conformation AP2σ2 R10 is in close contact with AP2β2 T9 and N10. The AP2σ2 R61 comes in close contact with AP2β2 K11 in the open conformation only, in which both R10 and R61 can be seen in close proximity to R15, which binds to the dileucine motif of the CaSR^1^.

HeLa cells were first confirmed to internalise Tf via CME that is dependent on AP2σ2, by showing that Alexa488-transferrin (Tf488) uptake was: detected in HeLa cells that had an acid wash to remove surface bound, non-internalized Tf488; blocked by Holotransferrin, a TfR binding competitor of Tf; and significantly reduced by siRNA targeting endogenous AP2σ2 (Fig. 2a). Next, Tf488 internalization was investigated in HeLa-AP2σ2H cells expressing the NDD-associated variants, and found to be reduced in R10W, R10Q, K18N and R61H cells, but not K18E cells, compared to WT (Fig. 2b). The FHH3-R15L mutant, which reduces CaSR signalling^1,4^, did not reduce Tf488 uptake (Fig. 2b), indicating that it did not affect TfR internalization, consistent with our previous observations in HEK293 cells^8^. We therefore hypothesised that NDD-associated AP2σ2 pathogenic variants are disrupting CME by possible effects on AP2μ2, which harbours the binding site for tyrosine-based motifs of cargo proteins, and that the underlying mechanisms may be due to AP2 complex conformational changes that result from disrupted interactions between the NDD-AP2σ2 variants and: other AP2σ2 residues; other AP2 complex subunits; the cargo protein; or intracellular proteins. To elucidate these mechanisms, we explored the effects of these NDD-AP2σ2 variants on the structure of the AP2 complex (Fig. 1) and undertook co-immunoprecipitation studies in HEK293-CF-AP2σ2H-WT and HEK293-CF-AσH-variant cells (Fig. 3 and Supplementary Fig. 5c-d).

**Figure 2.**
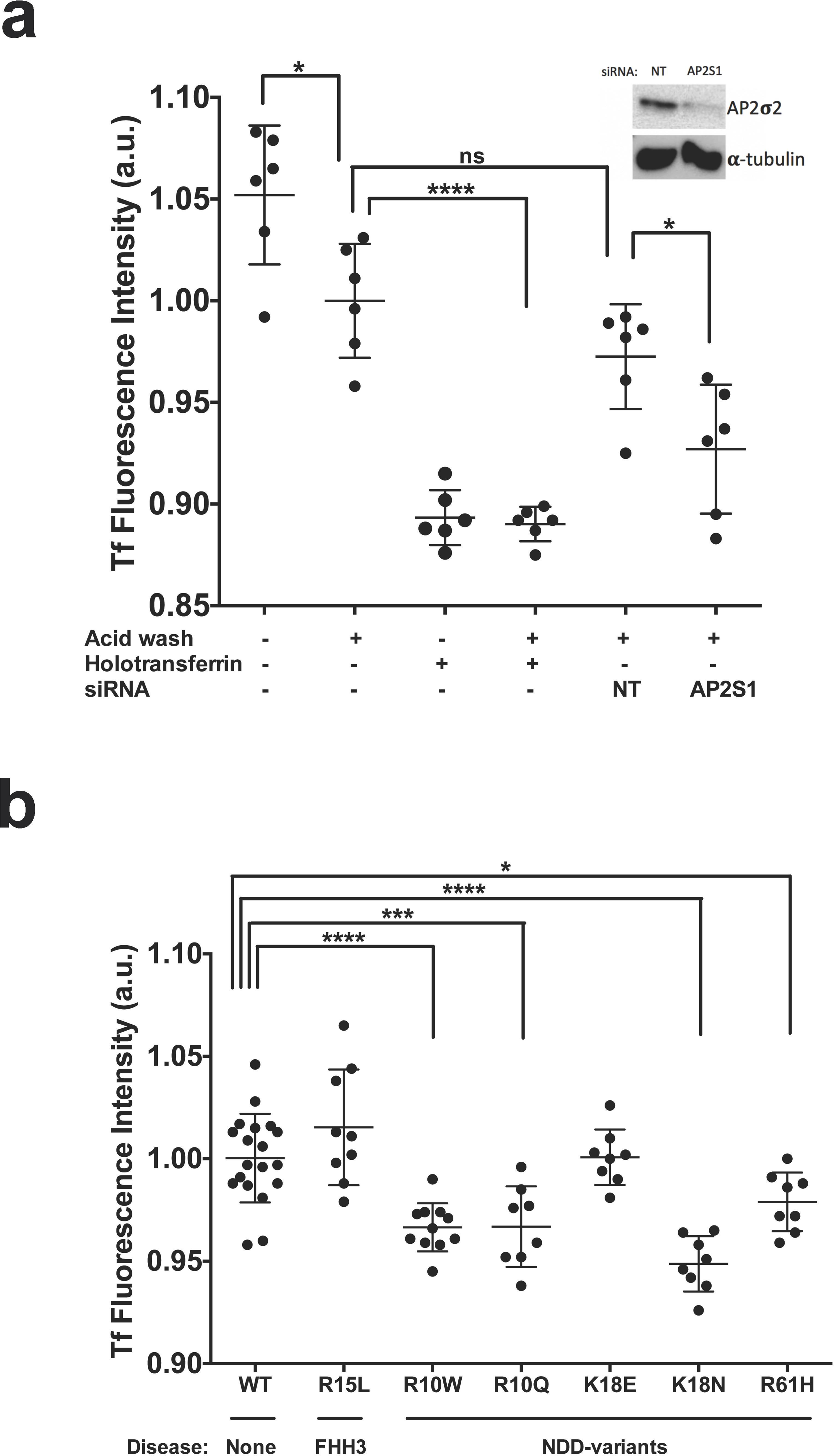
Transferrin-488 internalization in cells stably expressing AP2σ2-WT and - variant constructs. **a,** Transferrin (Tf)-488 uptake in HeLa cells is via transferrin receptors (TfRs) and dependent on AP2σ2 expression. The fluorescent signal from parental HeLa cells incubated with Tf488 is reduced following an acid wash due to the removal of surface bound Tf488 and therefore the fluorescent signal corresponds with the amount of internalized ligand. In cells treated with Holotransferrin competitor the fluorescent signal is near background, thereby confirming that Tf488 binding and internalization occurs via TfRs. Tf488 internalization is also reduced in cells treated with siRNA targeting *AP2S1* compared to non-targeting (NT) control siRNA, thereby confirming that AP2σ2 is involved in TfR uptake. Each bar represents the mean ± SD from 6 independent replicates. Data was analysed using an Ordinary One-Way ANOVA with Post-Hoc analysis performed using Tukey’s multiple comparisons test (selected comparisons are shown). Inset shows a Western blot of HeLa cells treated with *AP2S1* siRNA using anti-AP2σ2 antibodies confirming reduction of endogenous AP2σ2 expression. **b**, Tf488 internalization in HeLa cells stably expressing AP2σ2H-WT (WT) or NDD-AP2σ2H variants (-R10W, -R10Q, - R15L, -K18E, -K18N or -R61H), following cell incubation with Tf488, an acid wash to remove surface bound ligand, and measurement of the fluorescence signal. Tf488 internalization is reduced in cells expressing the NDD-AP2σ2H variants -R10W, -R10Q, - K18N and -R61H. Each bar represents the mean ± SD from 8-19 independent replicates. Tf internalization was compared to WT using ANOVA and Dunnett’s multiple comparisons test.

**Figure 3.**
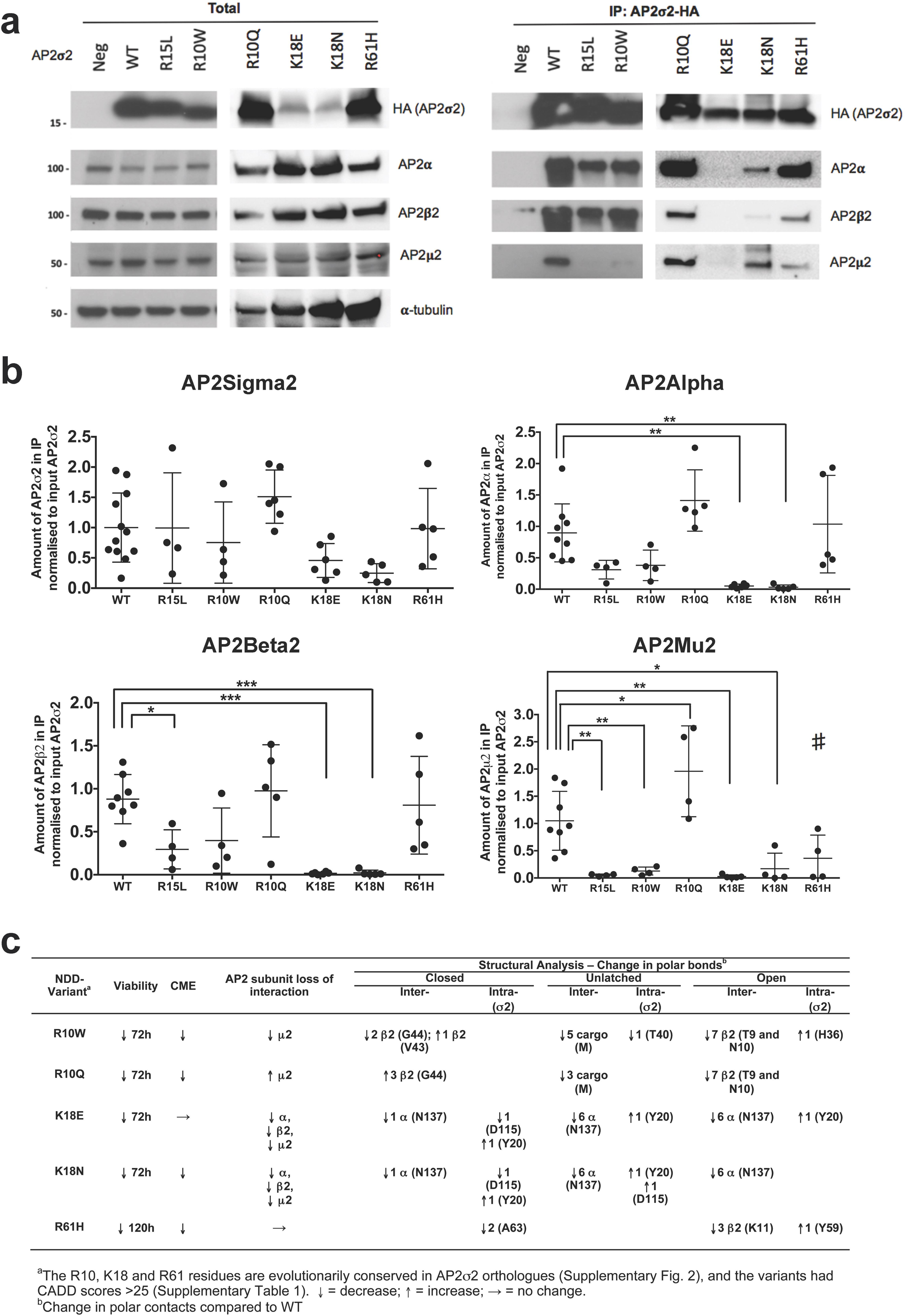
Co-immunoprecipitation showing AP2σ2 variants to have reduced interactions with AP2 subunits. **a,** Lysates of WT (HEK293-CF-AP2σ2H-WT) and NDD-patient associated variants (R10W, R10Q R15L, K18E, K18N, and R61H) or HEK293 only (Neg) cells underwent immunoprecipitation (IP) using anti-HA antibodies. Western blotting was performed on the input (Total) and the IP using anti-HA, anti-AP2α, anti-AP2β2, anti-Ap2µ2, and anti-α-tubulin antibodies. IP data shows AP2α and AP2β2 bands to be decreased in intensity in AP2σ2-K18E and AP2σ2-K18N cells, and AP2µ2 bands are decreased in AP2σ2 variants -R10W, -R15L, -K18E, -K18N and -R61H cells, when compared to AP2σ2-WT cells, suggesting reduced subunit interactions within the AP2 complex. **b**, Densitometry of Western blots (n = 4-12) to quantify AP2σ2, AP2α, AP2β2 and AP2µ2 in the IP of WT, and AP2σ2 variants (-R10W, -R10Q, -R15L, -K18E, -K18N and -R61H) cells, normalized to amount of AP2σ2 in the total lysate (pre-normalized to α-tubulin). Significantly reduced interactions are observed between: AP2µ2 and the AP2σ2 variants - R10W, -R10Q, -R15L, -K18E and -K18N; AP2α and the AP2⍰2 variants -K18E and - K18N; and AP2β2 and the AP2σ2 variants -R15L, -K18E and -K18N. Mean ± SD values are indicated and data were analyzed using an Ordinary One-Way ANOVA with Post-Hoc analysis performed using Dunnett’s multiple comparisons to WT test. *p<0.05, **p<0.01, ***p<0.001, #p=0.0719. **c**, Summary of cell viability, clathrin mediated endocytosis (CME), Co-IP and PyMOL analyses for the 5 NDD-AP2σ2 variants compared to WT.

Structural analysis using PyMOL of closed, unlatched and open AP2 conformations predicted each NDD-AP2σ2 variant to result in one or more disruptions in intra- and inter-subunit interactions, and loss of contacts with a cargo-motif peptide (Supplementary Fig. 6-8). For example, the AP2σ2-R10W and -R61H variants would result in loss or gain of intra-subunit polar contacts that potentially could alter structure of the AP2σ2 subunit, and also lead to reduced polar contacts with AP2β2; while the AP2σ2-K18E and -K18N variants would result in loss of polar contacts with AP2α. All of these would likely result in altered interactions between the NDD-AP2σ2 variants and other subunits that could impair the stability of the AP2 complex and inhibit the conformation change in the AP2μ2 subunit C-terminus, which is required to open and reveal the binding site for the tyrosine-based cargo-motif^3,10,11^. Co-immunoprecipitation studies confirmed these structural analysis predictions, by demonstrating 4 of the 5 NDD-associated AP2σ2 variants to have significantly altered interactions with Ap2μ2 (reduced for AP2σ2-R10W, -K18E, and - K18N; and increased for -R10Q), compared to WT (Fig. 3a-b). The NDD-associated AP2σ2-R61H variant also had reduced, but not statistically significant, interactions with Ap2μ2. The AP2σ2-K18E and -K18N variants additionally had decreased interactions with AP2α and AP2β2 subunits. These results indicate that the NDD-associated AP2σ2 variants are likely to impair stability of AP2 and hence CME (Fig. 3c). However, AP2σ2-K18E did not lead to decreased TfR uptake, and possible reasons may include: formation of very few faulty AP2 complexes with the observed TfR uptake resulting from AP2 complexes comprising only endogenous AP2σ2; or other mechanisms, e.g. loss of interactions with endocytic cargo-motifs or other intracellular proteins. Interestingly, the AP2σ2-R10 variants are predicted to lead to loss of polar contacts with the cargo peptide bearing the dileucine-motif (Supplementary Fig. 7), although a loss of interaction with CaSR, as seen with the AP2σ2 FHH3-R15L variant, was not observed (Supplementary Fig. 5c and d). This suggested that pathogenic mechanisms involving interactions with other intracellular proteins may be involved.

To investigate interactions of NDD-AP2σ2 variants with intracellular proteins, we focussed on the AP2σ2-R10W variant, as this was recurrent, identified in >75% of NDD-patients (Table 1), and consistently migrated further on Western blots than WT (Fig. 3a), thereby suggesting a reduced molecular weight. Thus, HA-tagged WT AP2σ2 had an apparent molecular weight of 18.4 kDa, whereas the AP2σ2-R10W mutant had an apparent molecular weight of ∼17 kDa (Fig. 4a). These differences were not due to protein folding, as they were observed on denaturing gels. We explored 3 other potential causes for this difference, namely splicing, post-translational modifications, or truncation. Use of prediction software (Fruitfly.org) indicated that *AP2S1* c.28C>T (GRCh38.p14; NM_004069.6; ENST00000263270.11), which is not near an intron-exon boundary, was unlikely to affect splicing. Post-translational modifications by methylation and glycosylation were excluded by: showing that treatment with an AMI-5 methylation inhibitor, or a PNGase enzyme, respectively, had no effect on AP2σ2 gel mobility (Fig. 4b-c): and proteomic analysis of immunoprecipitated AP2σ2-WT or AP2σ2-R10W proteins from HEK293-CF-AP2σ2H cells by liquid chromatography-tandem mass spectrometry (LC-MS/MS), which showed an absence of methylation, acetylation, phosphorylation, hydroxylation, ubiquitination, nitrosylation or lipidation abnormalities (Supplementary Fig. 9). C-terminal truncation appeared unlikely because of successful immunoprecipitation utilising the C-terminal HA-tag (Fig. 3a). We therefore explored the possibility of N-terminal truncation, by labelling immunoprecipitated AP2σ2-WT and AP2σ2-R10W with tandem mass tag zero (TMT0) using an N-terminal amine isotropic labelling of substrates (N-TAILS) strategy^14^, followed by digestion with trypsin and analysis by LC-MS/MS. Peptide fragments from the AP2σ2 N-terminal end spanning F4 to D26 were detected in both AP2σ2-WT and AP2σ2-R10W samples (Fig. 4d), thereby suggesting that an N-terminal truncation of AP2σ2 was not a plausible mechanism for the smaller mutant AP2σ2-R10W protein. However, in WT samples, a peptide with TMT0 label at F4 was detected with high confidence (Supplementary Table 2), while all peptide identifications covering F4 in the AP2σ2-R10W mutant were unlabelled (Fig. 4d, Supplementary Table 2), thereby suggesting that processing of the mutant protein may be altered, by an as yet to be identified mechanism.

**Figure 4.**
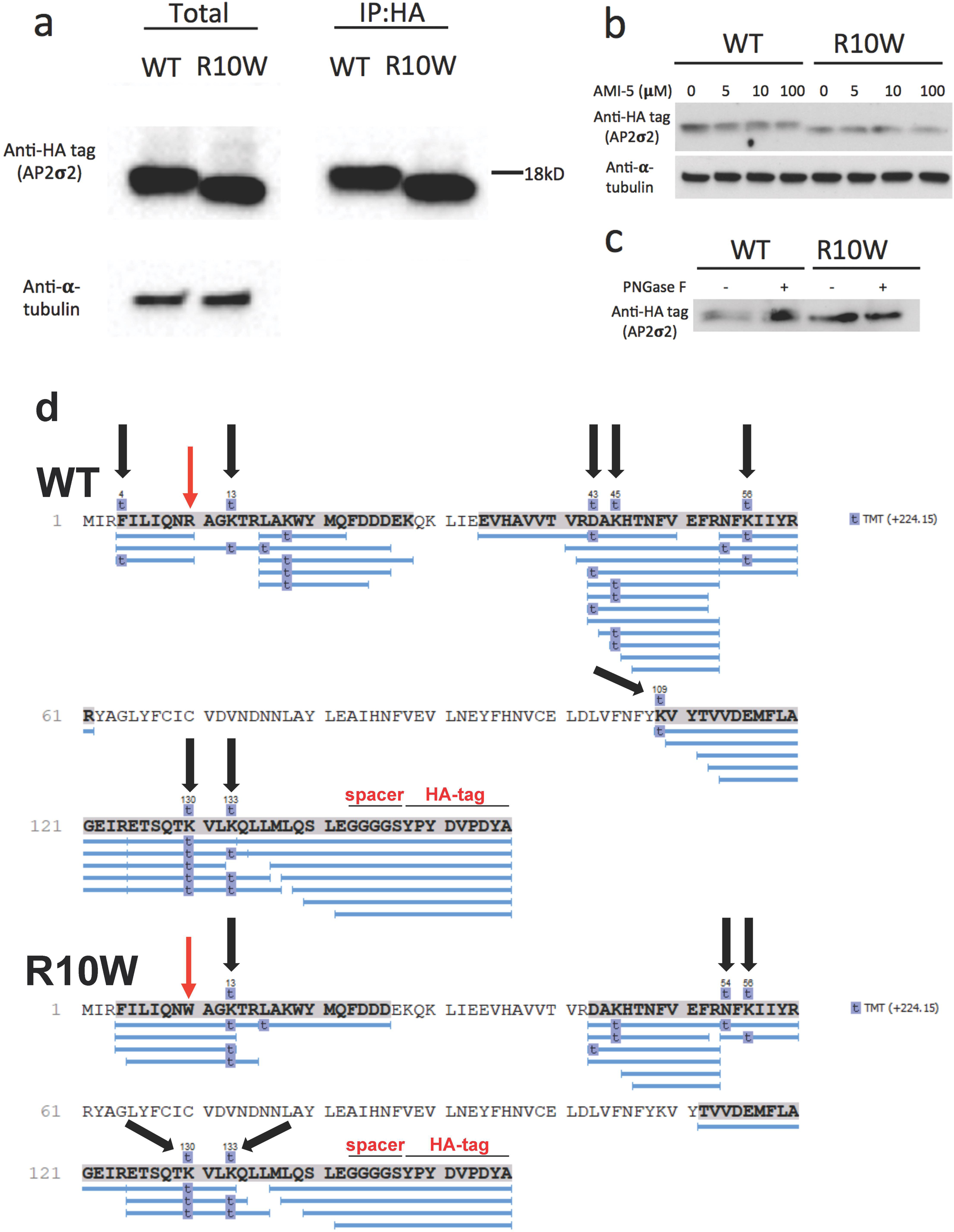
Investigation of mobility difference between AP2σ2-WT and variant AP2σ2-R10W by SDS PAGE. **a,** HA-tagged AP2σ2, which contains 156 amino acids (comprising 142 of AP2σ2, 5 of the GGGGS spacer and 9 of the HA tag), has a molecular weight of ∼18.4kDa. Western blot analysis of AP2σ2-R10W consistently showed it to have a reduced apparent molecular weight on a 4-20% gradient acrylamide gel compared to WT, when isolated directly from cells or following immunoprecipitation using anti-HA tag antibodies. **b**, Western blot analysis of HEK293 cells stably expressing FLAG-tagged CASR and HA-tagged WT AP2σ2 (HEK293-CF-AP2σ2H-WT) or -R10W treated with up to 100µM AMI-5 methylation inhibitor. Treatment did not alter the gel mobility of AP2σ2-WT or AP2σ2-R10W indicating that methylation is not causing the difference in molecular weight. **c**, Western blot analysis of immunoprecipitated AP2σ2 treated with 5 units of the deglycosylation enzyme PNGase F. Treatment did not alter the gel mobility of AP2σ2-WT or AP2σ2-R10W, indicating that glycosylation is not causing the difference in molecular weight. **d**, Protein N-terminal labelling with TMT0 analysis of immunoprecipitated AP2σ2-WT or AP2σ2-R10W, using MS. The amino acid sequence of AP2σ2-WT and AP2σ2-R10W is shown, with codon 10 indicated by the red arrow. The sequence starts at the N-terminus methionine (M) codon, and ends with a spacer (GGGGS) and HA tag (YPYDVPDYA) at the C-terminal end of the protein. Peptide coverage is denoted by the blue lines below the covered sequence shown in bold and highlighted in grey. Residues labelled with TMT0 (indicated by a t in a blue box) have a delta mass of +224.15. TMT0 will react with the N-termini of proteins as well as the free amino termini of lysine (K) residues. Residues shown with a TMT0 label above the sequence illustrated by the black arrows, indicate that the confidence filter for localisation (AScore >20) was met, whereas residues with TMT0 labels that are only below the sequence indicate that there was low localisation confidence at the spectrum level (see Supplementary Table 2).

To identify altered interactions of the AP2σ2-R10W variant with intracellular proteins, we analysed the AP2σ2 interactome. This revealed that 45 proteins were significantly (p<0.05) different in abundance between AP2σ2-WT and AP2σ2-R10W samples (Fig. 5a; Supplementary Table 3). STRING interaction analysis was used to generate a network of 43 of these proteins that were present in humans and in the STRING database, and this demonstrated a cluster of 8 proteins that were significantly less abundant in AP2σ2-R10W samples (Fig. 5b). These included AP2A1, AP2B1, AP3B1, ralA binding protein 1-associated Eps domain-containing 1 (REPS1), and Intersectin 1 (ITSN1) (Fig. 5b). Importantly, REPS1 and ITSN1 have known roles in neurodegeneration with brain iron accumulation due to abnormal TfR recycling^15^, and intellectual developmental delay^16^, respectively. We focussed further studies on ITSN1 as it is involved in: assembly and maturation of CCVs and synaptic vesicle recycling^17^; recruitment of dynamin and synaptojanin-1 to form clathrin coated pits^18,19^; endocytosis of membrane receptors including TfR^20^; synaptic vesicle endocytosis in brain neurones^21^ and interaction with REPS1^22^. Moreover, *ITSN1* deletions are associated with neurodevelopmental disorders^16,23,24^, while *Itsn1* knockout mice have abnormal synaptic vesicle recycling and learning deficits^25,26^. We therefore explored the AP2σ2-ITSN1 interaction by co-immunoprecipitation studies. ITSN1 has 13 and 17 isoforms in humans and mice respectively, with a ∼195kD isoform most highly expressed in human and mouse brain, while a ∼137kD isoform is detected in multiple tissues including brain and skeletal muscle (GTEx Analysis Release V8). Co-immunoprecipitation analysis using normal mouse brain, revealed that while the 195kD ITSN1 isoform was highly expressed, it was the ∼137kD isoform that predominantly co-immunoprecipitated with AP2σ2 (Fig. 5c). Moreover, analysis of the AP2σ2-ITSN1 interaction in HEK293-AP2σ2H-WT and HEK293-AP2σ2H-R10W cells revealed that the ∼137kD ITSN1 isoform was significantly (p<0.05) decreased in the IP from AP2σ2-R10W, compared to AP2σ2-WT cells (Fig. 5d-e), thereby validating the reduced interaction observed in the AP2σ2 interactome data.

**Figure 5.**
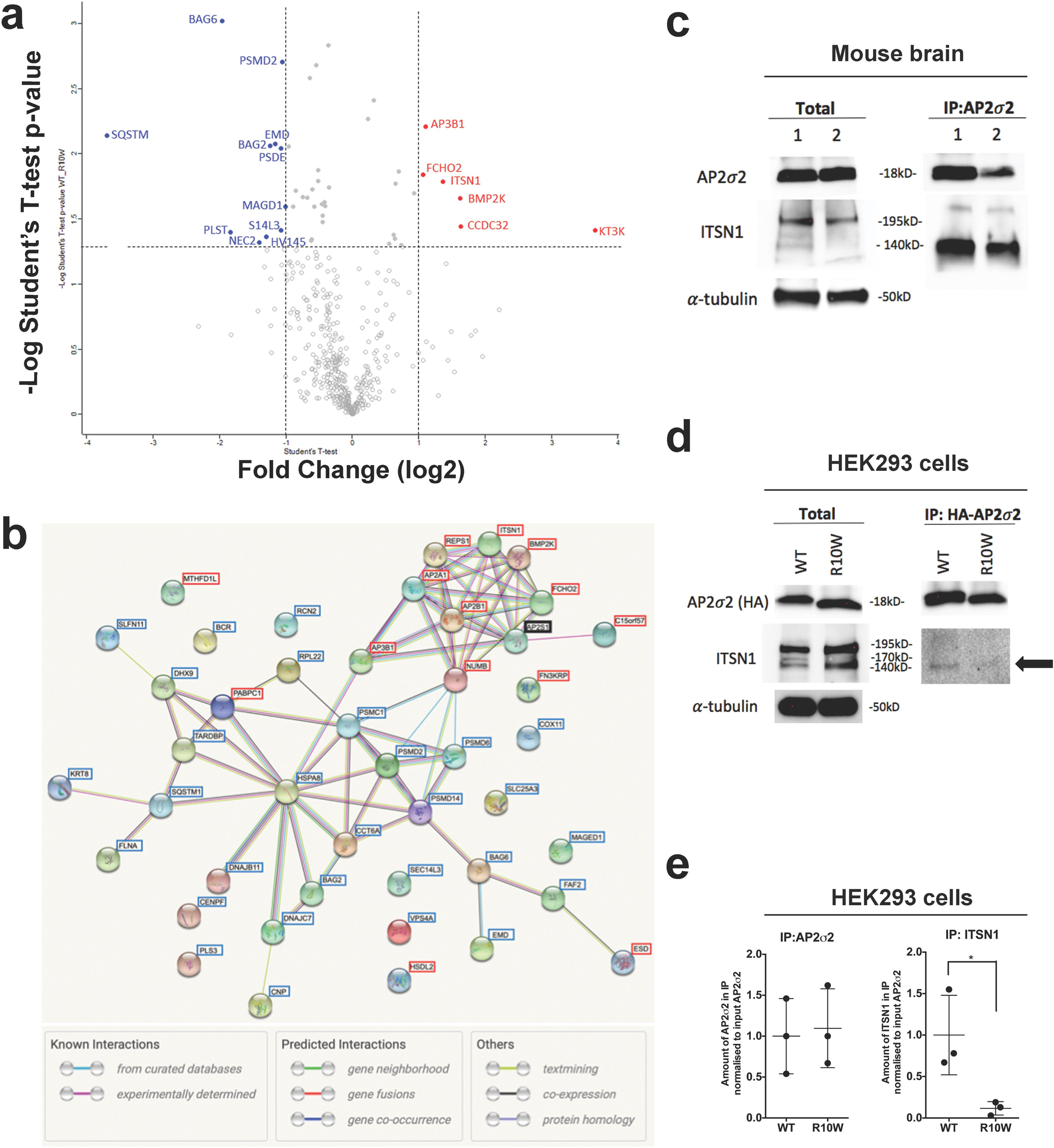
*AP2S1* STRING Interactome analysis. Lysates from HEK-293 cells stably expressing AP2σ2-WT or NDD-associated AP2σ2-R10W variant underwent immunoprecipitation using anti-HA antibodies, and the immunoprecipitated proteins were digested with elastase and analysed by LC-MS/MS. Forty-five proteins that showed a significant (p<0.05) difference in abundance between WT and R10W samples were identified (shown in Supplementary Table 3). **a,** Volcano plot comparing the interactome of AP2σ2-WT versus variant AP2σ2-R10W expressing cells. The fold change in protein levels (in log 2) is displayed on the x-axis and the -log10 t-test p-value on the y-axis. Proteins with p-values <0.05 (log p>1.301) are shown with filled circles and coloured in when quantified with a fold change difference greater than 2 (blue higher in AP2σ2-R10W and red higher in AP2σ2-WT). **b,** The interactions of AP2σ2, encoded by the *AP2S1* gene (shown with a black border), with 43 of the 45 proteins were analysed using the STRING database of known and predicted protein-protein interactions^39,40^. The 2 proteins excluded from the STRING analysis were: the human herpesvirus type 7 strain JI nuclear egress protein 2 (NEC2), because it is not a human protein; and the Immunoglobulin heavy variable 1-45 (IGHV1-45) protein because it was not present in the STRING database. Genes encoding proteins that were more abundant in AP2σ2-WT samples compared to AP2σ2-R10W samples are shown with a red border, while proteins that were more abundant in the AP2σ2-R10W samples compared to AP2σ2-WT samples are shown with a blue border. Network statistics: number of nodes = 44; number of edges = 74; average node degree = 3.36; average local clustering coefficient 0.48; expected number of edges = 21; PPI enrichment p-value <1.0e-16. A cluster of 8 proteins that interact with AP2σ2 are shown in the top right corner and comprised AP3 subunit beta-1 (AP3B1); AP2 subunit alpha-1 (AP2A1); AP2 subunit beta-1 (AP2B1); RalA binding protein 1-associated Eps domain containing protein 1 (REPS1); Intersectin 1 (ITSN1); Bone morphogenic protein-2 inducible protein kinase (BMP2K); Fes/CIP4 homology and mu domain containing endocytic adaptor 2 (FCHO2); and Coiled coil domain-containing 32 (CCDC32, also known as C15orf57). LC-MS/MS analysis revealed that the AP2σ2-R10W variant has a reduced interaction with ITSN1 (Supplementary Table 3). To validate this finding co-immunoprecipitation studies were performed in normal mouse brain and human HEK293 cells expressing AP2σ2-WT or variant AP2σ2-R10W, as follows. **c**, Lysates of B6J-H normal mouse brain (n=2) were co-immunoprecipitated (IP) using anti-AP2σ2 antibodies and Western blotting performed on the input (Total) and the IP using anti-AP2σ2, anti-ITSN1, and anti-α-tubulin antibodies. IP data shows the longer 195kD ITSN1 isoform to be most highly expressed in the total murine brain lysate, but the shorter ∼137kD isoform is predominantly co-immunoprecipitated with AP2σ2. **d**, Lysates of HEK293-CF-AP2σ2H-WT or -R10W cells co-immunoprecipitated using anti-HA antibodies and Western blotting performed on the input (Total) and the IP using anti-HA, anti-ITSN1, and anti-α-tubulin antibodies. Western blot data suggests that in AP2σ2-WT and AP2σ2-R10W cells, the ∼195kD and ∼137kD ITSN1 isoforms are expressed at similar levels, but the shorter ∼137kD ITSN1 isoform is detected in the AP2σ2-WT IP, which is consistent with data observed in the murine brain. In contrast, this interaction is lost (arrow) in the AP2σ2-R10W variant cells, thereby indicating a reduced interaction between variant AP2σ2 and ITSN1. **e**, Densitometry of Western blots (n = 3) to quantify AP2σ2 and ITSN1 in the IP of AP2σ2-WT and AP2σ2-R10W cells, normalized to amount of AP2σ2 in the total lysate (pre-normalized to α-tubulin). A significantly reduced interaction is confirmed between AP2σ2-R10W and ITSN1. Mean ± SD values are indicated and data was analyzed using an unpaired t-test. *p<0.05.

Thus, AP2σ2 variants such as R10W, R10Q, K18E, K18N and R61H, have been shown to affect CME and/or interactions with other AP2 subunits or proteins involved in vesicle recycling. Our findings that the AP2σ2 R10W variant disrupts the AP2σ2-ITSN1 interaction highlights a potential mechanism for altering synaptic function that results in NDD and epilepsy in patients. Furthermore, AP2σ2 pathogenic variants may be more common than previously thought in causing NDD, as *AP2S1* was reported to be among 72 and 285 genes that were significantly associated with developmental disorders following analysis of exome sequencing data from 20,627 individuals with autism spectrum disorder and 31,058 parent-offspring trios of individuals with developmental delay, respectively^27,28^. Defects in CME are also associated with cognitive impairment in human disorders, e.g. Alzheimer’s disease and Down Syndrome^29–33^, and as a disease mechanism for NDD and generalized epilepsy (MIM ⋕618587) in 4 patients with a missense (p.Arg170Trp) variant of AP2µ2^34^.

In summary, we have demonstrated that AP2σ2 variants associated with NDD and epilepsy have reduced interactions with AP2 subunits, that can lead to decreases in cell viability and CME uptake of TfR, and in the case of AP2σ2-R10W reduced interactions with multiple proteins including ITSN1, which may affect brain development and physiology that leads to cognitive defects and epileptic seizures.

## Data Availability

All data produced in the present study are available upon reasonable request to the authors

## Methods

Methods are available online.

## Acknowledgements

This work was supported by a Wellcome Trust Senior Clinical Investigator Award (RVT), and National Institute for Health Research (NIHR) – Oxford Biomedical Research Centre Programme grant. RVT is a Wellcome Trust Investigator and NIHR Senior Investigator. For the purpose of Open Access, the author has applied a CC BY public copyright licence to any Author Accepted Manuscript (AAM) version arising from this submission. We are grateful to the patients and families that participated in this research. The authors are grateful to the participants of the MyCode Community Health Initiative for the use of their genomic and electronic health information. The patient enrolment and exome sequencing for the DiscovEHR study were funded by the Regeneron Genetics Center. We would like to acknowledge the Geisinger-Regeneron DiscovEHR Collaboration for making the genotype data and phenotype available for this project. This research was also made possible through access to data in the National Genomic Research Library, which is managed by Genomics England Limited (a wholly owned company of the Department of Health and Social Care). The National Genomic Research Library holds data provided by patients and collected by the NHS as part of their care and data collected as part of their participation in research. The National Genomic Research Library is funded by the National Institute for Health Research and NHS England. The Wellcome Trust, Cancer Research UK and the Medical Research Council have also funded research infrastructure. Mass spectrometry analysis was performed at the Discovery Proteomics Facility (headed by Roman Fischer) which is part of the TDI MS Laboratory (led by Benedikt Kessler). KGC is Chairholder of the Emil von Behring Chair for Neuromuscular and Neurodegenerative Disorders by CSL Behring. KGC is member of the European Reference Network for Rare Neuromuscular Diseases (ERN EURO-NMD) and of the European Reference Network for Rare Neurological Diseases (ERN-RND). KGC reports no disclosures relevant to the manuscript.

## Author Contributions

M.S, A.L.B and R.V.T. designed the experiments. M.S., A.L.B., V.J.S., K.E., K.G.K., K.E.L., and R.V.T. performed the experiments and analysed the data. M.S., A.L.B. R.H., I.V. and R.F. conducted the LC/MS/MS experiments and analysed data. N.dL. and K.L.I.V.G performed genetic analysis. M.E.W.A.A., M.B., A. Be., A.Bo., R.B., D.J.C., D.A.C., A.C., K.G.C., B.C., G.C., A.S.D.P., E.J.D., E.D., D.A.D., B.G., T.B.H., J.S.H., S.H., R.A.H., B.I., L.I., A.J., J.J., J.J.K., B.K., E.W.K., E.K., B.C.L., E.L.M., E.J.M., K.M., B.A.M., A.M., M.O., A.P., A.R., V.R., P.R., S.S., A.L.S., D.T.S., L.S.B., F.T.M.T., C.T., A.T., K.V.S., G.V., M.V.L., M.W., M.H.W., L.C.W.K., A.W., I.Z., G.E.R.C. and R.V.T. recruited patients. M.S., F.M.H. and R.V.T. drafted the manuscript. All authors critically evaluated the manuscript.

## On line Methods

The methods include details on: patients and ethical approval; DNA sequence analysis; generation and maintenance of cell lines; cell viability assay; transferrin assay; in silico analysis; mouse study approval; immunoprecipitation and western blot analysis; post-translational modification assays; proteomic analysis (N-TAILS); proteomic analysis (AP2σ2 interactome); and statistical analysis.

### Patients and Ethical approvals

Informed consent was obtained from individuals or their family/guardians/parents, using protocols approved by the local and national ethics committees. Unrelated individuals and their parents with *AP2S1* p.Arg10Trp (n=20), p.Arg10Gln (n=3), p.Lys18Glu (n=1), p.Lys18Asn (n=1) and p.Arg61His (n=1) variants were investigated (Table 1).

### DNA Sequence Analysis

Leukocyte DNA was used with *AP2S1*-specific primers for PCR amplification, and the DNA sequences of the PCR products were determined using the Big Dye Terminator v1.1 cycle sequencing kit (Life Technologies) and an ABI3130 automated capillary sequencer (Applied Biosystems). A representative DNA sequence chromatogram indicating a heterozygous C-to-T transition (CGG to TGG) at nucleotide c.28 in exon 2 of *AP2S1* (NM_004069.6) is shown for patient ID7 (Supplementary Fig. 1)

### Generation and maintenance of cell lines

HeLa cell lines were maintained in DMEM media supplemented with 10% FCS. HeLa cells stably expressing WT (HeLa-AP2σ2H-WT) or variant (HeLaAP2σ2H -R10W, HeLa-AP2σ2H-R10Q, HeLa-AP2σ2H-R15L, HeLa-AP2σ2H-K18E, HeLa-AP2σ2H-K18N, and HeLa-AP2σ2H-R61H) AP2σ2 proteins, were generated using a pcDNA5 construct, containing a full-length human *AP2S1* cDNA^8^, with a C-terminal HA tag (YPYDVPDYA), and maintained under hygromycin selection. Site directed mutagenesis using the Q5 Site-Directed Mutagenesis kit (New England Biolabs) and *AP2S1* specific primers (Thermo Fisher Scientific) were used to generate the mutant *AP2S1* constructs. Western blotting of cell lysates using anti-HA tag antibodies was performed to confirm AP2σ2-HA expression (Supplementary Fig. 3). Human embryonic kidney (HEK293) cell lines were maintained in DMEM media supplemented with 10% foetal calf serum (FCS). HEK293 cells stably expressing the CaSR bearing an N-terminal FLAG tag (HEK293-CF), were generated as previously described^41^. Briefly a pcDNA3 construct (Invitrogen) containing a full length human *CASR* cDNA^42^ with an N-terminal FLAG tag (DYKDDDDK) was used to generate HEK293-CF cells and stably maintained under G418 (geneticin) selection. HEK293-CF cells stably expressing WT (HEK293-CF-AP2σ2H-WT) or variant (HEK293-CF-AP2σ2H-R10W, HEK293-CF-AP2σ2H -R10Q, HEK293-CF-AP2σ2H-R15L, HEK293-CF-AP2σ2H-K18E, HEK293-CF-AP2σ2H-K18N and HEK293-CF-AP2σ2H-R61H) AP2σ2 proteins, were generated using the pcDNA5 construct described above, and maintained under hygromycin and geneticin selection. Serum response element (SRE), and nuclear factor of activated T-cells (NFAT) intracellular calcium mobilization assays were performed using methods previously described^43,44^ in HEK293-CF cells transiently transfected with HA-tagged *AP2S1* WT or mutant expression constructs.

### Cell Viability Assays

Cell viability was assessed using the CellTiter Blue Cell Viability assay (Promega), whereby 20 µL of CellTiter Blue reagent was added per well, incubated for 1 h at 37°C, 5% (vol/vol) CO_2_ and the fluorescent outputs read on a CytoFluor microplate reader (PerSeptive Biosystems, MA, USA) at 530 nm excitation and 580 nm emission^45^.

### Transferrin Assay

HeLa, HeLa-AP2σ2H-WT, HeLa-AP2σ2H-R10W, HeLa-AP2σ2H-R10Q, HeLa-AP2σ2H-R15L, HeLa-AP2σ2H-K18E, HeLa-AP2σ2H-K18N, and HeLa-AP2σ2H-R61H cells were seeded in 96-well plates and cultured for 24 h. Cells treated with siRNA were incubated with 50nM non-targeting (NT) control (ON-Targetplus Control Pool, Dharmacon) or *AP2S1* siRNA (ONTargetplus Human *AP2S1* specifically targeting the 3’UTR only, Dharmacon) complexed with jetPRIME according to the manufacturer’s guidelines (Polyplus), and cultured for 48 h. Cells were then placed in basal media (DMEM, 0.1%BSA, 20mM HEPES) for 45 min at 37°C, and incubated with 25μg/ml Transferrin-AlexFluor488 conjugate (Thermo Scientific) on ice for 30 min. Control cells were co-incubated with 250μg/ml Holotransferrin (Sigma). Cells were next washed in ice-cold Dulbecco’s Phosphate-Buffered Saline (DPBS; Sigma) and incubated in pre-warmed basal media for 15 min to allow Tf488 internalisation. After this cells were washed once in ice-cold DPBS, followed by three washes in ice-cold DPBS or acid (0.2N acetic acid, 150mM NaCl, pH2.7) for 3 min each. Next, the cells were washed in DPBS and fixed in 4% paraformaldehyde for 15 min on ice, after which they were washed once more in DPBS and fluorescent intensity analysed using a PHERAstar platereader (BMG Labtech). Measurements were taken using an optic module set to a wavelength of 485-520nm. Results were analysed using PHERAstar FS MARS software and readings normalised using mock treated cells that were not treated with Tf488.

### In silico analyses

Protein sequences of AP2σ2 orthologues and paralogues were analysed with Homologene (https://www.ncbi.nlm.nih.gov/homologene). The crystal structure of the AP2 heterotetramer in the closed unbound conformation (PDB entry 2VGL)^3^, the unlatched conformation in association with a RM(phosphoS)QIKRLLSE peptide from CD4 representing the dileucine-based [ED]xxxL[LI] endocytic cargo motif (PDB 2JKR)^10^, and the open conformation in association with a tyrosine-based YxxΦ motif containing peptide (DYQRLN) from TGN38 but no [ED]xxxL[LI] peptide (PDB 2XA7)^11^, have been reported. The PyMOL Molecular Graphics System (version 2.4.0, Schrodinger) was used to model the effect of the Arg10Trp, Arg10Gln, Lys18Glu, Lys18Asn, and Arg61His variants on the interactions with the AP2 complex using the three-dimensional structure of AP2 archived in the Protein Data Bank at the European Bioinformatics Institute with the accession numbers 2VGL, 2JKR and 2XA7. The splice site prediction tool Fruitfly.org was used to assess changes to splicing patterns within the *AP2S1* gene relating to the c.28C>T variant. AP2σ2 interacting proteins identified in the Proteomic analysis as being significantly different in abundance between WT and R10W samples were analysed using the STRING Protein-Protein Interaction Networks Functional Enrichment Analysis tool maintained by the Swiss Institute of Bioinformatics, CPR Novo Nordisk Foundation Centre Protein Research and European Molecular Biology Laboratory^39,40^.

### Mouse Study Approval

All animal studies were approved by the Medical Research Council Harwell Institute Ethical Review Committee and were licenced under the Animal (Scientific Procedures) Act 1986, issued by the UK Government Home Office Department (PPL PP9613750).

### Immunoprecipitation and Western Blot Analysis

Cells were lysed in ice-cold lysis buffer (0.5% NP40, 135mM NaCl, 20mM Tris pH7.5, 1mM EDTA, 1x protease inhibitor (Sigma-Aldrich) 2mM Na_3_VO_4_, 10mM NaF) and debris cleared by centrifugation. Immunoprecipitations using anti-CASR antibodies, were performed by mixing lysates with antibody for 30 min at 4°C prior to addition to protein G agarose beads (Cell Signalling Technology) and further mixing for 2h. Alternatively, lysates were mixed with anti-FLAG-sepharose or anti-HA-sepharose beads (Cell Signalling Technology) and mixed for 2h. Beads were then washed 5x with lysis buffer, eluted with FLAG or HA peptide as appropriate, and proteins prepared in 4x Laemmli loading dye containing β-mercaptoethanol and resolved using 4-15% or 4-20% SDS-PAGE gel electrophoresis. Proteins were transferred to polyvinylidene difluoride membrane and probed with primary antibodies (CASR (ADD, Abcam), AP2σ2 (Ab128950, Abcam), FLAG (ab49763, Abcam), HA (CST3724, Cell Signalling Technologies), AP2α (BD610501, BD Biosciences), AP2β2 (BD610381, BD Biosciences), AP2µ2 (Ab137727, Abcam), ITSN1 (NBPI-71832, Novus Biologicals) and α-tubulin (Ab15246, Abcam), and horse radish peroxidase (HRP)-conjugated secondary antibodies (715-035-150 and 711-035-152, Jackson ImmunoResearch), prior to visualisation using Pierce ECL Western blotting substrate. Alpha-tubulin protein expression was used as a loading control. Densitometry analysis was performed by calculating the number of pixels per band using ImageJ software. Whole brains that had been snap frozen in liquid nitrogen from B6J-H mice were obtained. Frozen brain tissue was crushed using a mortar and pestle and resuspended in ice-cold lysis buffer for 20 min and centrifuged to clear debris. To preclear the lysate, Protein A agarose beads (Cell Signalling Technology) were added and mixed for 1 h at 4°C. Beads were then spun and the supernatant recovered and added to anti-AP2σ2 antibodies which were then mixed on a rotator for 30 min at 4°C. The antibody containing lysate was then added to pre-washed Protein A agarose beads and incubated overnight at 4°C. Beads were then washed and immunoprecipitated proteins recovered in 4x Laemmli loading dye containing β-mercaptoethanol and resolved using 4-20% SDS-PAGE gel electrophoresis.

### Post-translational Modification Assays

Methylation of AP2σ2 was investigated using an AMI-5 methylation inhibitor (Abcam). HEK293-CF-AP2σ2H-WT and HEK293-CF-AP2σ2H-R10W cells were seeded into 24-well plates and cultured for 24 h. Cells were then treated with 0, 5μM, 10μM or 100μM AMI-5 methylation inhibitor (Abcam) and cultured for a further 24 h. Cells were then lysed in ice-cold lysis buffer and lysates resolved using 4-15% SDS-PAGE gel electrophoresis analysed by Western blotting using anti-HA and anti-α-tubulin antibodies. Glycosylation of AP2σ2 was investigated using PNGase F to digest immunoprecipitated AP2σ2 protein. HEK293-CF-AP2σ2H-WT and HEK293-CF-AP2σ2H-R10W cell lysates were immunoprecipitated as described above using anti-HA-sepharose beads, and AP2σ2 eluted using 1mg/ml HA peptide (Sino Biological) in phosphate buffered saline (PBS) for 15min at 37°C. The eluted protein was then incubated with 5 units of PNGase F (Sigma) for 3h at 37°C, resolved using 4-20% SDS-PAGE gel electrophoresis and analysed by Western blotting using anti-HA antibodies.

### Proteomic Analysis - N-Terminal Amine Isotropic labelling of Substrates (N-TAILS) Tandem Mass Tag Zero (TMT0) Labelling of AP2_t7_2 proteoforms

HEK293-CF-AP2σ2H-WT and HEK293-CF-AP2σ2H-R10W cells were lysed in ice cold buffer and IPs performed by mixing the lysates with anti-HA-sepharose beads for 48 h at 4°C. Beads were washed 3x with lysis buffer, buffer removed and proteins labelled using 0.8mg Tandem Mass Tag zero (TMT0; Thermo Fisher Scientific) resuspended in anhydrous acetonitrile for 2 h. Tris buffered saline (TBS) was added to quench the reaction and beads spun down and washed. Supernatant was removed and protein eluted using 4x Laemmli loading dye containing β-mercaptoethanol and resolved using 4-20% SDS-PAGE gel electrophoresis. Samples were treated either by Western blot analysis using anti-HA antibodies, or by silver staining using Pierce silver stain kit according to the manufacturer’s instructions. Silver stained bands of 16-19kDa were excised from the gel, destained using Proteosilver destained kit and then digested with trypsin following the standard in-gel-digest approach. Samples were then analysed by liquid chromatography-tandem mass spectrometry (LC-MS/MS) using the Dionex Ultimate 3000 Ultra Performance Liquid Chromatography (UPLC) coupled to a TimsTof Pro mass spectrometer (BrukerDaltonics). Briefly, peptides were loaded into an IonOptiks column (75 µm x 150 mm C18 column with 1.6 µm particles) at a flow rate of 400nl/min and analysed using a 47 min linear gradient from 2 to 35% solvent B (A: 0.1% formic acid in water; B: 0.1% FA in acetonitrile). MS data were acquired in TimsTof Pro operated in PASEF mode. The ion mobility window was set to 1/k0 start = 0.6 Vs/cm2 to 1/k0 end = 1.6 Vs/cm2, ramp time 166 ms with locked duty cycle, mass range 100 - 1700 m/z. MS/MS were acquired in 10 PASEF frames (1.89 sec cycle). Target intensity was set to 20000 and threshold intensity 1000. MS raw files were analysed using PEAKSX (Bioinformatics solutions Inc). Data were searched against the human UniProtKB/Swiss-Prot database (downloaded on the 20^th^ September 2020 containing 23155 sequences) containing the HA-tag AP2σ2 WT and mutant (R10W) sequences, using a 20ppm peptide and 0.04 Da MS/MS mass tolerance, respectively; selecting trypsin as enzyme with a semi-specific mode (3 missed cleavages allowed); carbamidomethylation (C) as variable modification, and oxidation (M), deaminadation (NQ) and TMT0 as variable modification in addition to a PEAKS-PTM search (1% false discovery rate (FDR) at peptide spectrum match (PSM) level).

### Proteomic Analysis - AP2σ2 Interactome

HEK293-CF-AP2σ2H-WT or HEK293-CF-AP2σ2H-R10W cells were lysed in ice cold buffer and IPs performed using anti-HA-sepharose as described above (n=3). For on-bead digest, beads were washed with PBS and buffer removed before freezing them at - 80°C. Beads were resuspended in 400mM Tris-HCl and digested overnight with 1ug of elastase at 37°C. Peptides were desalted by solid phase extraction using SOLA HRP PE cartridges (ThermoFischer) and dried down. Dried peptides were re-constituted in LC-MS grade water containing 2% acetonitrile and 0.1% Trifluoroacetic acid (TFA) and analysed by LC_MS/MS using a Dionex Ultimate 3000 UPLC coupled to a Q-Exactive classic spectrometer (Thermo Fisher Scientific). In brief, peptides were first loaded onto a trap column (PepMapC18; 300µm x 5mm, 5µm particle size, Thermo Fischer), separated on an EasySpray column (50cm; ES803, Thermo Fischer) over a 60 min gradient of 2 to 35% acetonitrile in 0.1% formic acid and 5% DMSO with a flow rate of 250 nL/min, and analysed in a Q-Exactive operated in a data-dependent acquisition mode (DDA). Full MS scans were acquired in the Orbitrap with 70k resolution (AGC target at 3e6 ions) and scan range between 380 – 1800 m/z. Top fifteen precursor ions (charge state >+2) were sequentially isolated in the Quad (1.6 m/z window), fragmented on the HCD cell (NCE of 28%) and analysed in the orbitrap with a resolution of 17.5K (128ms maximum acquisition time; AGC target of 1e5 and 27 sec dynamic exclusion). The AP2σ2 interactome was analysed using the ProgenesisQI (v4.1, Non-linear dynamics, Waters) label free quantitation platform combined with PEAKSX (Bionformatics solutions Inc.) data search engine. In brief, Progenesis peaks list was searched against the Human UniprotSwissprot database containing the HA-tag AP2σ2 WT and R10W sequences. Search parameters included a 10ppm peptide and 0.05Da MS/MS mass tolerance, respectively, no enzyme (for elastase), carboamidomethylation (C) as fixed modification and Oxidation (M) and Deaminidation (NQ) as variable modifications. A 1% FDR at PSM level was applied to the PEAKS-PTM output and the pep.xml output was imported back to ProgenesisQI. Data were normalised to all proteins identified and quantified using HiN relative quantitation (topN 3). ANOVA p values were determined with p<0.05 classified as significant. The Progenesis output was further analysed using Perseus (v1.6.2.2). Briefly, normalised protein abundances were log 2 transformed and missing values were imputed following normal distribution. A two-sample student t-test combined with permutation FDR (5%) was performed. Student t-test difference and -10log student t-test were used to generate a scatter plot. The mass spectrometry raw data included in this paper had been deposited to the Proteome eXchange Consortium via the PRIDE partner repository with the dataset identifier PXDXXXXX^46^.

### Statistical Analysis

Densitometry of co-immunoprecipitation assays and fluorescent intensity from the Tf488 uptake assays were analysed using an Ordinary One-Way ANOVA with Post-Hoc analysis performed using Tukey’s multiple comparisons test or Dunnett’s multiple comparisons test as stated. All analyses were performed using GraphPad Prism (GraphPad), and a value of P<0.05 was considered significant.

