## Supplementary Figure legends for "Adaptor protein 2 sigma subunit (*AP2S1*) variants associated with neurodevelopmental disorders"

**Supplementary Figure 1**. **DNA sequencing confirms AP2S1 mutation a**. DNA sequence analysis showing c.28C>T (highlighted) within exon 2 of *AP2S1* (numbering begins from ATG). The chromatograms show that the patient ID7 (Table 1) with neurodevelopmental delay (NDD) is heterozygous C/T, while both parents are homozygous C/C. **b**. The AP2S1 c.28C>T substitution is predicted to lead to a missense substitution of Arg (R), encoded by CGG, to Trp (W), encoded by TGG, at codon 10. **c**. Pedigree showing affected proband is heterozygous (C/T) at c.28, while both parents are homozygous (C/C).

**Supplementary Figure 2**. **Evolutionary conservation of the AP2σ2 Arg10, Lys18 and Arg61 variants. a,** Multiple protein sequence alignments of AP2σ2 revealed evolutionary conservation of Arg10 (R10), Lys18 (K18) and Arg61 (R61) residues (indicated with arrows) in orthologues and human paralogues. Conserved residues are shaded grey. The R10, K18 and R61 residues are conserved in all AP2σ2 orthologues. There is also conservation of R10 and R61 in AP1 subunit paralogues, and K18 in AP1, AP3S1 and AP4 subunit paralogues. The highly conserved Arg15 (R15) residue mutated in patients with FHH3 is indicated with an asterisk (*).

**Supplementary Figure 3**. **AP2σ2-HA stable expression in HeLa cells (HeLa-AP2σ2H).** Western blotting of HeLa cell lysates using anti-HA antibodies confirms AP2σ2-HA expression in the cells stably transfected with AP2σ2 wild-type (WT) or variant (-R10W, -R10Q, -R15L, -K18E, -K18N and -R61H) constructs.

**Supplementary Figure 4. Relative cell viability of HeLa-AP2σ2H-WT, and HeLa-AP2σ2H-R61H cell lines after 120h.** The viability rates were normalized such that WT cells reached 1.0 after 72h. Error bars represent SD. The data was analysed using an unpaired t-test. ***p<0.001.

**Supplementary Figure 5. CaSR signaling and interactions with WT AP2σ2, FHH3-associated R15L AP2σ2, and NDD-associated R10W AP2σ2.** HEK293 cells stably expressing the FLAG-tagged CaSR (HEK293-CF) were transfected with HA-tagged *AP2S1* (WT or variant (R15L or R10W) plasmid DNA and CaSR signaling assessed by **a**, Extracellular calcium ($Ca^{2+}_e$)-induced NFAT luciferase reporter responses (n=4), and **b**, Extracellular calcium ($Ca^{2+}_e$)-induced SRE luciferase reporter responses (n=4). There are no significant differences in NFAT or SRE responses between cells expressing AP2**σ**2 WT and R10W. **c**, Lysates from the cells underwent IP using anti-FLAG antibodies. Western blotting was performed on the input (Total) and the IP using anti-FLAG, anti-HA**,** and anti-**α**-tubulin antibodies. The IP data shows that the AP2σ2 band is reduced in intensity in the R15L cells compared to WT indicating a reduced interaction with CaSR as previously reported[41]. In contrast, there is no reduction in the AP2**σ** band from the R10W cells, indicating no loss of interaction with the CaSR. **d**, Densitometry of the Western blotting (n = 4-8) to quantify AP2σ2 in the IP of WT, and variant cells, normalized to the amount of CASR in the total lysate (pre-normalized to α-tubulin). The FHH3-AP2**σ**2 R15L had a reduction in the interaction with CaSR, compared to AP2σ2-WT and the NDD-R10W variant, whereas the interactions of the AP2σ2-WT and NDD-R10W variants with the CaSR were not significantly (NS) different. Mean

± SD values are indicated and were compared to WT using ANOVA and Dunnett's multiple comparisons test. *p<0.05.

**Supplementary Figure 6**. **PyMOL analysis of the polar contacts involving residues 10, 18 and 61 of the AP2σ2 subunit in the closed AP2 conformation.** Images of the AP2 tetramer shown in the closed conformation (Protein Data Bank entry 2VGL[3]) with regions of AP2α (green), AP2β2 (blue), AP2σ2 (grey) and AP2μ2 (yellow) illustrated; only the side chains of specific residues are shown for simplicity. Residues 10, 18 and 61 are shown in red and residue 15 is shown in orange. Polar contacts from these specific residues within AP2σ2 (intrachain), or between other subunits (interchain), are shown as blue or black dotted lines, respectively. **a**, AP2σ2 Arg10 (R10) is predicted to have two interchain polar contacts with AP2β2 Gly44 (G44) in addition to an intrachain polar contact with AP2σ2 Gly64 (G64). **b**, AP2σ2 Trp10 (W10) loses the two polar contacts with AP2β2 G44, but acquires a single intrachain contact with AP2β2 Val43 (V43). **c**, AP2σ2 Gln10 (Q10) is predicted to increase the number of interchain polar contacts with AP2β2 G44 to 5. **d**, AP2σ2 K18 has an interchain polar contact with AP2α Asn137 (N137), in addition to intrachain polar contacts with AP2σ2 Ile5 (I5) and Asp115 (D115). **e**, AP2σ2 Asn18 (N18) loses the interchain contact with AP2α N137 but acquires a new intrachain contact with AP2σ2 Tyr20 (Y20) which replaces the contact lost with AP2σ2 D115. **f**, AP2σ2 Glu18 (E18) also loses contact with the AP2α N137 but acquires a new intrachain contact with AP2σ2 Tyr20 (Y20) which replaces the contact lost with AP2σ2 D115. **g**, AP2σ2 Arg61 (R61) has two intrachain polar contacts with AP2σ2 Ala63 (A63). **h**, AP2σ2 His61 (H61) lose the contacts with AP2σ2 A63.

**Supplementary Figure 7**. **PyMOL analysis of the polar contacts involving residues 10, 18 and 61 of the AP2σ2 subunit in the unlatched AP2 conformation.** PyMOL analysis of the AP2 complex in the unlatched conformation (UC) with an RM(phosphoS)QIKRLLSE peptide from CD4 representing the dileucine-based [ED]xxxL[LI] endocytic cargo motif ((Protein Data Bank (PDB) entry 2JKR[16]). The AP2 complex subunits AP2α (green), AP2β2 (blue), AP2σ2 (grey) and AP2μ2 (yellow), together with the RM(phosphoS)QIKRLLSE cargo peptide (pink), are shown; only the side chains of specific residues are shown for simplicity. Residues 10, 18 and 61 are shown in red, while the predicted polar contacts from these specific residues within AP2σ2 (intrachain), or between other subunits or cargo peptide (interchain), are shown as blue or black dotted lines, respectively. **a**, AP2σ2 Arg10 (R10) has 5 points of contact with the Met (M) residue of the RM(phosphoS)QIKRLLSE cargo peptide (ranging in distance from 3.3 to 4.0A), in addition to intrachain polar contacts with AP2σ2 Thr40 (T40) and Gly64 (G64). AP2σ2 Arg15 (R15; shown in orange) has an interchain contact (3.7A) with the Gln (Q) residue of the cargo peptide, as previously reported[1]. **b**, AP2σ2 variant Trp10 (W10) has lost all contacts with M on the cargo peptide, but acquired a new polar contact with AP2σ2 Ala63 (A63) which replaces one with AP2σ2 G64. **c**, AP2σ2 variant Gln10 (Q10) loses 3 of the 5 polar contacts with M on the cargo peptide. **d**, AP2σ2 Lys18 (K18) has 6 interchain polar contacts with AP2α Asn137 (N137), and 2 intrachain polar contacts with AP2σ2 Ile5 (I5). **e**, The AP2σ2 variant Glu18 (E18) has lost all the interchain contacts with AP2α N137 but acquired a new intrachain polar contact with AP2σ2 Y20. **f,** AP2σ2 variant Asn18 (N18) loses all interchain contacts with AP2α N137, but acquires new intrachain contacts with AP2σ2 Tyr20 (Y20) and Asp115 (D115). **g**, AP2σ2 Arg61 (R61) has no polar contacts. **h**, The AP2σ2 variant His61 (H61) has no polar contacts.

**Supplementary Figure 8. PyMOL analysis of the polar contacts involving residues 10, 18 and 61 of the AP2σ2 subunit in the open AP2 conformation.** PyMOL analysis of the AP2 complex in the open conformation (OC) with a DYQRLN peptide representing the tyrosine-based YxxΦ endocytic cargo motif (PDB entry 2XA7[17]) (not shown). In the open conformation the AP2μ2 subunit has been repositioned due to the large conformational change within the AP2 complex. The AP2 complex subunits AP2α (green), AP2β2 (blue), AP2σ2 (grey) and AP2μ2 (yellow) are shown; only the side chains of specific residues are shown for simplicity. Residues 10, 18 and 61 are shown in red, while the predicted polar contacts from these specific residues within AP2σ2 (intrachain), or between other subunits (interchain), are shown as blue or black dotted lines, respectively. **a**, AP2σ2 R10 has 15 interchain polar contacts with AP2β2 via the Thr9 (T9) and Asn10 (N10) residues, and an intrachain polar contact with AP2σ2 G64. **b**, AP2σ2 W10 loses 7 of the 15 interchain polar contacts with the AP2β2 T9 and N10 residues, but acquires an additional intrachain contact with AP2σ2 His36 (H36). **c**, AP2σ2 Q10 also loses 7 of the 15 interchain polar contacts with the AP2β2 T9 and N10 residues. **d**, AP2σ2 K18 has 6 points of interchain contact with AP2α N137, in addition to 2 intrachain polar contacts with AP2σ2 I5. **e**, AP2σ2 E18 loses all
