## Supplementary Table 1 for "Adaptor protein 2 sigma subunit (*AP2S1*) variants associated with neurodevelopmental disorders"

| AP2S1 Variant | REVEL score | CADD score | Source |
|---|---|---|---|
| R10W | 0.471 | 32.0 | GeneDx |
| R10Q | 0.273 | 28.5 | GeneDx, DiscovEHR |
| K18E | 0.699 | 30.0 | Genomics England |
| K18N | 0.673 | 27.0 | GeneDx |
| R61H | 0.447 | 29.8 | GeneDx |

**Supplementary Table 1.** Computational analysis of AP2$\sigma$2 variants using Rare Exome Variant Ensemble Learner (REVEL) and Combined Annotation Dependent Deletion (CADD). REVEL scores are based on a combination of scores from 13 tools (MutPred, FATHMM v2.3, VEST 3.0, PolyPhen-2, SIFT, PROVEAN, Mutation Assesor, MutationTaster, LRT, GERP++, SiPhy, phyloP and phastCons) (Ioannidis et al. Am. J. Hum. Genet. 2016 99:877-885). Scores range from 0 to 1 with higher scores reflecting a greater likelihood that the variant is disease-causing. 75.4% of disease mutations but only 10.9% of neutral variants have a REVEL score >0.5. CADD scores are based on genomic features from surrounding sequence context, gene model annotations, evolutionary constraint, epigenetic measurements and functional predictions (Kircher et al. 2014 Nat. Genet 46:310-315). The higher the CADD score the more the variant is predicted to be deleterious. Thus, variants with CADD scores >10, >20 and >30 are predicted to be among the 10%, 1% and 0.1% most deleterious possible substitutions in the human genome.
