## Supplementary Table 2 for "Adaptor protein 2 sigma subunit (*AP2S1*) variants associated with neurodevelopmental disorders"

| AP2S1 WT | | | | | | | | | | | |
|---|---|---|---|---|---|---|---|---|---|---|---|
| **Protein Accession** | **Peptide** | **Start** | **End** | **-10lgP** | **Mass** | **Length** | **ppm** | **m/z** | **z** | **PTM** | **AScore** |
| Q00000\|HA-AP2S1 WT_HUMAN | R.F(+224.15)ILIQNR.A | 4 | 10 | 36.06 | 1126.686 | 7 | -0.1 | 564.3503 | 2 | TMT | F1:TMT:1000.00 |
| Q00000\|HA-AP2S1 WT_HUMAN | R.FILIQNRAGK(+224.15)TR.L | 4 | 15 | 25.31 | 1639.989 | 12 | -1.3 | 547.6694 | 3 | TMT | K10:TMT:62.82 |
| Q00000\|HA-AP2S1 WT_HUMAN | R.LAK(+224.15)WYMQFDDD.E | 16 | 26 | 56.57 | 1654.77 | 11 | -2.4 | 828.3904 | 2 | TMT | K3:TMT:11.10 |
| Q00000\|HA-AP2S1 WT_HUMAN | R.L(+224.15)AKWYM(+15.99)QFDDD.E | 16 | 26 | 46.53 | 1670.765 | 11 | -1.2 | 836.3888 | 2 | TMT; Oxidation (M) | L1:TMT:0.00;M6:Oxidation (M):1000.00 |
| Q00000\|HA-AP2S1 WT_HUMAN | R.D(+224.15)AKHTNFVEFR.N | 43 | 53 | 73.05 | 1586.82 | 11 | 0.1 | 529.9474 | 3 | TMT | D1:TMT:22.85 |
| Q00000\|HA-AP2S1 WT_HUMAN | R.D(+224.15)AKHTNFVEF.R | 43 | 52 | 34.02 | 1430.719 | 10 | -3.5 | 716.3644 | 2 | TMT | D1:TMT:8.22 |
| Q00000\|HA-AP2S1 WT_HUMAN | R.DAK(+224.15)HTN(+.98)FVEFR.N | 43 | 53 | 66.38 | 1587.804 | 11 | -3.1 | 530.2738 | 3 | TMT; Deamidation (NQ) | K3:TMT:14.04;N6:Deamidation (NQ):1000.00 |
| Q00000\|HA-AP2S1 WT_HUMAN | R.NFK(+224.15)IIYR.R | 54 | 60 | 38.06 | 1176.702 | 7 | -1.6 | 589.3573 | 2 | TMT | K3:TMT:39.76 |
| Q00000\|HA-AP2S1 WT_HUMAN | R.N(+.98)(+224.15)FKIIYR.R | 54 | 60 | 25.57 | 1177.686 | 7 | -1.7 | 589.8492 | 2 | Deamidation (NQ); TMT | N1:Deamidation (NQ):1000.00;N1:TMT:0.00 |
| Q00000\|HA-AP2S1 WT_HUMAN | R.ETSQTK(+224.15)VLK(+224.15)QLL.M | 125 | 136 | 45.16 | 1835.113 | 12 | 1 | 612.7123 | 3 | TMT | K6:TMT:39.44;K9:TMT:77.91 |
| Q00000\|HA-AP2S1 WT_HUMAN | R.ETSQTK(+224.15)VLK.Q | 125 | 133 | 46.18 | 1256.734 | 9 | 0.3 | 629.3745 | 2 | TMT | K6:TMT:22.69 |

| AP2S1 R10W | | | | | | | | | | | |
|---|---|---|---|---|---|---|---|---|---|---|---|
| **Protein Accession** | **Peptide** | **Start** | **End** | **-10lgP** | **Mass** | **Length** | **ppm** | **m/z** | **z** | **PTM** | **AScore** |
| Q00001\|HA-AP2S1 R10W_HUMAN | R.FILIQNWAGK(+224.15)TR.L | 4 | 15 | 58.93 | 1669.967 | 12 | -1.1 | 835.9897 | 2 | TMT | K10:TMT:181.94 |
| Q00001\|HA-AP2S1 R10W_HUMAN | R.L(+224.15)AKWYMQFDDD.E | 16 | 26 | 43.37 | 1654.77 | 11 | 0.5 | 828.3928 | 2 | TMT | L1:TMT:0.00 |
| Q00001\|HA-AP2S1 R10W_HUMAN | R.DAK(+224.15)HT(+79.96)NFVEFR.N | 43 | 53 | 74.34 | 1666.777 | 11 | 0.9 | 556.6002 | 3 | TMT; Sulfation | K3:TMT:8.22;T5:Sulfation:1000.00 |
| Q00001\|HA-AP2S1 R10W_HUMAN | R.D(+224.15)AKHT(+79.96)NFVEFR.N | 43 | 53 | 70.15 | 1666.777 | 11 | 0.9 | 556.6002 | 3 | TMT; Sulfation | D1:TMT:0.00;T5:Sulfation:1000.00 |
| Q00001\|HA-AP2S1 R10W_HUMAN | R.D(+224.15)AKHTNFVEFR.N | 43 | 53 | 68.29 | 1586.82 | 11 | -0.4 | 529.9472 | 3 | TMT | D1:TMT:14.04 |
| Q00001\|HA-AP2S1 R10W_HUMAN | R.DAK(+224.15)HTN(+.98)FVEFR.N | 43 | 53 | 42.32 | 1587.804 | 11 | 3.4 | 530.2772 | 3 | TMT; Deamidation (NQ) | K3:TMT:0.00;N6:Deamidation (NQ):1000.00 |
| Q00001\|HA-AP2S1 R10W_HUMAN | R.N(+224.15)FKIIYR.R | 54 | 60 | 22.03 | 1176.702 | 7 | -4.9 | 393.2393 | 3 | TMT | N1:TMT:24.32 |
| Q00001\|HA-AP2S1 R10W_HUMAN | R.NFK(+224.15)IIYR.R | 54 | 60 | 35.87 | 1176.702 | 7 | -0.9 | 589.3577 | 2 | TMT | K3:TMT:36.05 |
| Q00001\|HA-AP2S1 R10W_HUMAN | R.ETSQTK(+224.15)VLK(+224.15)QLL.M | 125 | 136 | 39.05 | 1835.113 | 12 | 3.8 | 612.714 | 3 | TMT | K6:TMT:45.16;K9:TMT:81.34 |

**Supplementary Table 2.** Table indicating the AScore (confidence in localisation) of all the peptides that were identified in carrying a TMT0 label. Unhighlighted rows indicate peptides containing TMT0 labelled lysine residues. Yellow highlighted rows indicate non-lysine TMT0 labelled residues that have an AScore >20 and thus meet the confidence level set for localisation; such residues could represent N-termini. Grey highlighted rows indicate non-lysine TMT0 labelled residues that have an AScore <20 and thus there is not enough evidence in terms of the number of MS fragment ions for the software to confidently assign the TMT0 label to the residue.
