## Supplementary Table 3 for "Adaptor protein 2 sigma subunit (*AP2S1*) variants associated with neurodevelopmental disorders"

| Accession | Peptide count | Unique peptides | Anova (p) | Max fold change | Highest Mean Condition | Lowest Mean Condition | Description |
|---|---|---|---|---|---|---|---|
| P46379\|BAG6_HUMAN | 3 | 3 | 0.00095 | 3.854 | R10W | WT | Large proline-rich protein BAG6 GN=BAG6 |
| P35268\|RL22_HUMAN | 3 | 3 | 0.00146 | 1.281 | R10W | WT | 60S ribosomal protein L22 GN=RPL22 |
| Q13200\|PSMD2_HUMAN | 5 | 5 | 0.00197 | 2.064 | R10W | WT | 26S proteasome non-ATPase regulatory subunit 2 GN=PSMD2 |
| P11274\|BCR_HUMAN | 1 | 1 | 0.00210 | 1.458 | R10W | WT | Breakpoint cluster region protein GN=BCR |
| Q15008\|PSMD6_HUMAN | 2 | 2 | 0.00261 | 1.562 | R10W | WT | 26S proteasome non-ATPase regulatory subunit 6 GN=PSMD6 |
| Q6UB35\|C1TM_HUMAN | 2 | 2 | 0.00390 | 1.252 | WT | R10W | Monofunctional C1-tetrahydrofolate synthase, mitochondrial GN=MTHFD1L |
| P11940\|PABP1_HUMAN | 3 | 3 | 0.00539 | 1.177 | WT | R10W | Polyadenylate-binding protein 1 GN=PABPC1 |
| O00203\|AP3B1_HUMAN | 1 | 1 | 0.00619 | 2.168 | WT | R10W | AP-3 complex subunit beta-1 GN=AP3B1 |
| Q13501\|SQSTM_HUMAN | 1 | 1 | 0.00721 | 13.257 | R10W | WT | Sequestosome-1 GN=SQSTM1 |
| P50402\|EMD_HUMAN | 2 | 2 | 0.00843 | 2.223 | R10W | WT | Emerin GN=EMD |
| O95816\|BAG2_HUMAN | 6 | 6 | 0.00869 | 2.353 | R10W | WT | BAG family molecular chaperone regulator 2 GN=BAG2 |
| P05787\|K2C8_HUMAN | 10 | 7 | 0.00882 | 1.958 | R10W | WT | Keratin, type II cytoskeletal 8 GN=KRT8 |
| O00487\|PSDE_HUMAN | 2 | 2 | 0.00904 | 2.061 | R10W | WT | 26S proteasome non-ATPase regulatory subunit 14 GN=PSMD14 |
| Q9UN37\|VPS4A_HUMAN | 3 | 3 | 0.01335 | 1.428 | R10W | WT | Vacuolar protein sorting-associated protein 4A GN=VPS4A |
| O95782\|AP2A1_HUMAN | 154 | 103 | 0.01362 | 1.642 | WT | R10W | AP-2 complex subunit alpha-1 GN=AP2A1 |
| Q0JRZ9\|FCHO2_HUMAN | 11 | 11 | 0.01444 | 2.070 | WT | R10W | F-BAR domain only protein 2 GN=FCHO2 |
| P11142\|HSP7C_HUMAN | 75 | 27 | 0.01621 | 1.421 | R10W | WT | Heat shock cognate 71 kDa protein GN=HSPA8 |
| Q15811\|ITSN1_HUMAN | 4 | 4 | 0.01646 | 2.564 | WT | R10W | Intersectin-1 GN=ITSN1 |
| P10768\|ESTD_HUMAN | 34 | 33 | 0.01695 | 1.557 | WT | R10W | S-formylglutathione hydrolase GN=ESD |
| P21333\|FLNA_HUMAN | 41 | 41 | 0.01810 | 1.273 | R10W | WT | Filamin-A GN=FLNA |
| Q9UBS4\|DJB11_HUMAN | 5 | 5 | 0.01877 | 1.534 | R10W | WT | DnaJ homolog subfamily B member 11 GN=DNAJB11 |
| Q13148\|TADBP_HUMAN | 1 | 1 | 0.01917 | 1.791 | R10W | WT | TAR DNA-binding protein 43 GN=TARDBP |
| Q96D71\|REPS1_HUMAN | 15 | 15 | 0.02002 | 1.914 | WT | R10W | RalBP1-associated Eps domain-containing protein 1 GN=REPS1 |
| Q14257\|RCN2_HUMAN | 16 | 16 | 0.02159 | 1.677 | R10W | WT | Reticulocalbin-2 GN=RCN2 |
| Q08211\|DHX9_HUMAN | 2 | 2 | 0.02177 | 1.586 | R10W | WT | ATP-dependent RNA helicase A GN=DHX9 |
| Q9NSY1\|BMP2K_HUMAN | 31 | 29 | 0.02204 | 3.251 | WT | R10W | BMP-2-inducible protein kinase GN=BMP2K |
| Q99615\|DNJC7_HUMAN | 4 | 4 | 0.02349 | 1.342 | R10W | WT | DnaJ homolog subfamily C member 7 GN=DNAJC7 |
| P09543\|CN37_HUMAN | 1 | 1 | 0.02424 | 1.367 | R10W | WT | 2',3'-cyclic-nucleotide 3'-phosphodiesterase GN=CNP |
| P40227\|TCPZ_HUMAN | 5 | 5 | 0.02523 | 1.327 | R10W | WT | T-complex protein 1 subunit zeta GN=CCT6A |
| Q9Y5V3\|MAGD1_HUMAN | 1 | 1 | 0.02531 | 1.985 | R10W | WT | Melanoma-associated antigen D1 GN=MAGED1 |
| Q96CS3\|FAF2_HUMAN | 5 | 5 | 0.02531 | 1.817 | R10W | WT | FAS-associated factor 2 GN=FAF2 |
| P62191\|PRS4_HUMAN | 3 | 2 | 0.02558 | 1.482 | R10W | WT | 26S proteasome regulatory subunit 4 GN=PSMC1 |
| Q7Z7L1\|SLN11_HUMAN | 7 | 6 | 0.02982 | 1.370 | R10W | WT | Schlafen family member 11 GN=SLFN11 |
| P49454\|CENPF_HUMAN | 1 | 1 | 0.03358 | 1.358 | R10W | WT | Centromere protein F GN=CENPF |
| Q9BV29\|CCD32_HUMAN | 3 | 3 | 0.03632 | 3.171 | WT | R10W | Coiled-coil domain-containing protein 32 GN=CCDC32 |
| Q9UDX4\|S14L3_HUMAN | 1 | 1 | 0.03874 | 2.040 | R10W | WT | SEC14-like protein 3 GN=SEC14L3 |
| Q9HA64\|KT3K_HUMAN | 2 | 2 | 0.03887 | 7.488 | WT | R10W | Ketosamine-3-kinase GN=FN3KRP |
| P13797\|PLST_HUMAN | 1 | 1 | 0.04000 | 3.248 | R10W | WT | Plastin-3 GN=PLS3 |
| P49757\|NUMB_HUMAN | 4 | 4 | 0.04198 | 1.521 | WT | R10W | Protein numb homolog GN=NUMB |
| A0A0A0MS14\|HV145_HUMAN;A0A0B4J2H0\|HV69D_HUMAN;A0A0C4DH29\|HV103_HUMAN;A0A0C4DH33\|HV124_HUMAN;A0A0C4DH39\|HV158_HUMAN;A0A0G2JMI3\|HV692_HUMAN;P01742\|HV169_HUMAN;P01743\|HV146_HUMAN;P0DP01\|HV108_HUMAN | 1 | 1 | 0.04328 | 2.488 | R10W | WT | Immunoglobulin heavy variable 1-45 GN=IGHV1-45 |
| P63010\|AP2B1_HUMAN | 232 | 174 | 0.04467 | 1.533 | WT | R10W | AP-2 complex subunit beta GN=AP2B1 |
| Q00325\|MPCP_HUMAN | 7 | 7 | 0.04519 | 1.554 | R10W | WT | Phosphate carrier protein, mitochondrial GN=SLC25A3 |
| Q9Y6N1\|COX11_HUMAN | 1 | 1 | 0.04663 | 1.553 | R10W | WT | Cytochrome c oxidase assembly protein COX11, mitochondrial GN=COX11 |
| P52466\|NEC2_HHV7J | 1 | 1 | 0.04817 | 2.765 | R10W | WT | Nuclear egress protein 2 OS=Human herpesvirus 7 (strain JI) OX=57278 GN=NEC2 |
| Q6YN16\|HSDL2_HUMAN | 3 | 3 | 0.04897 | 1.486 | WT | R10W | Hydroxysteroid dehydrogenase-like protein 2 GN=HSDL2 |
| Q00000\|HA-AP2_HUMAN | 35 | 3 | 0.04993 | 1.732 | WT | R10W | GN=HA-AP2 |
| Q00001\|HA-AP2-R10W_HUMAN | 34 | 2 | 0.04993 | 1.732 | WT | R10W | GN=HA-AP2 |
| P63104\|1433Z_HUMAN | 7 | 3 | 0.05176 | 1.649 | WT | R10W | 14-3-3 protein zeta/delta GN=YWHAZ |
| P62333\|PRS10_HUMAN | 2 | 1 | 0.05425 | 2.042 | R10W | WT | 26S proteasome regulatory subunit 10B GN=PSMC6 |
| P17980\|PRS6A_HUMAN | 1 | 1 | 0.05480 | 2.283 | R10W | WT | 26S proteasome regulatory subunit 6A GN=PSMC3 |
| O75643\|U520_HUMAN | 1 | 1 | 0.05523 | 1.839 | R10W | WT | U5 small nuclear ribonucleoprotein 200 kDa helicase GN=SNRNP200 |
| P60953\|CDC42_HUMAN | 1 | 1 | 0.05704 | 1.279 | WT | R10W | Cell division control protein 42 homolog GN=CDC42 |
| P04844\|RPN2_HUMAN | 3 | 3 | 0.05714 | 1.444 | R10W | WT | Dolichyl-diphosphooligosaccharide--protein glycosyltransferase subunit 2 GN=RPN2 |
| P12236\|ADT3_HUMAN | 17 | 1 | 0.05999 | 1.465 | R10W | WT | ADP/ATP translocase 3 GN=SLC25A6 |

| | | | | | | | |
|---|---|---|---|---|---|---|---|
| P49368\|TCPG_HUMAN | 9 | 8 | 0.06274 | 1.622 | R10W | WT | T-complex protein 1 subunit gamma GN=CCT3 |
| P62195\|PRS8_HUMAN | 5 | 4 | 0.06291 | 1.282 | R10W | WT | 26S proteasome regulatory subunit 8 GN=PSMC5 |
| O75815\|BCAR3_HUMAN | 2 | 2 | 0.06397 | 1.491 | R10W | WT | Breast cancer anti-estrogen resistance protein 3 GN=BCAR3 |
| O43242\|PSMD3_HUMAN | 4 | 4 | 0.06727 | 1.979 | R10W | WT | 26S proteasome non-ATPase regulatory subunit 3 GN=PSMD3 |
| O14950\|ML12B_HUMAN;P19105\|ML12A_HUMAN | 2 | 2 | 0.06874 | 1.943 | R10W | WT | Myosin regulatory light chain 12B GN=MYL12B |
| P30041\|PRDX6_HUMAN | 1 | 1 | 0.06930 | 1.681 | R10W | WT | Peroxiredoxin-6 GN=PRDX6 |
| Q86X55\|CARM1_HUMAN | 1 | 1 | 0.06970 | 2.395 | WT | R10W | Histone-arginine methyltransferase CARM1 GN=CARM1 |
| Q7Z6Z7\|HUWE1_HUMAN | 18 | 18 | 0.07107 | 1.989 | R10W | WT | E3 ubiquitin-protein ligase HUWE1 GN=HUWE1 |
| P21964\|COMT_HUMAN | 1 | 1 | 0.07158 | 1.393 | R10W | WT | Catechol O-methyltransferase GN=COMT |
| Q7L8L6\|FAKD5_HUMAN | 1 | 1 | 0.07261 | 1.648 | R10W | WT | FAST kinase domain-containing protein 5, mitochondrial GN=FASTKD5 |
| P35637\|FUS_HUMAN | 1 | 1 | 0.07319 | 1.986 | R10W | WT | RNA-binding protein FUS GN=FUS |
| Q9P2R7\|SUCB1_HUMAN | 3 | 3 | 0.07385 | 1.435 | R10W | WT | Succinate--CoA ligase [ADP-forming] subunit beta, mitochondrial GN=SUCLA2 |
| Q9H4B7\|TBB1_HUMAN | 47 | 1 | 0.07407 | 2.625 | R10W | WT | Tubulin beta-1 chain GN=TUBB1 |
| P17812\|PYRG1_HUMAN | 4 | 4 | 0.07632 | 1.073 | WT | R10W | CTP synthase 1 GN=CTPS1 |
| P49411\|EFTU_HUMAN | 34 | 34 | 0.07817 | 1.200 | R10W | WT | Elongation factor Tu, mitochondrial GN=TUFM |
| P50990\|TCPQ_HUMAN | 23 | 22 | 0.07886 | 1.435 | WT | R10W | T-complex protein 1 subunit theta GN=CCT8 |
| Q99460\|PSMD1_HUMAN | 3 | 3 | 0.07975 | 1.293 | R10W | WT | 26S proteasome non-ATPase regulatory subunit 1 GN=PSMD1 |
| Q6PD74\|AAGAB_HUMAN | 34 | 34 | 0.08130 | 1.294 | WT | R10W | Alpha- and gamma-adaptin-binding protein p34 GN=AAGAB |
| P01889\|HLAB_HUMAN;P17693\|HLAG_HUMAN | 1 | 1 | 0.08157 | 1.768 | R10W | WT | HLA class I histocompatibility antigen, B alpha chain GN=HLA-B |
| P24311\|COX7B_HUMAN | 1 | 1 | 0.08199 | 1.675 | R10W | WT | Cytochrome c oxidase subunit 7B, mitochondrial GN=COX7B |
| A6NEC2\|PSAL_HUMAN;P55786\|PSA_HUMAN | 1 | 1 | 0.08402 | 2.076 | R10W | WT | Puromycin-sensitive aminopeptidase-like protein GN=NPEPPSL1 |
| Q9BS26\|ERP44_HUMAN | 1 | 1 | 0.08722 | 1.169 | R10W | WT | Endoplasmic reticulum resident protein 44 GN=ERP44 |
| P24534\|EF1B_HUMAN | 7 | 6 | 0.09054 | 2.003 | R10W | WT | Elongation factor 1-beta GN=EEF1B2 |
| Q07666\|KHDR1_HUMAN | 1 | 1 | 0.09144 | 2.171 | R10W | WT | KH domain-containing, RNA-binding, signal transduction-associated protein 1 GN=KHDRBS1 |
| Q9NVZ3\|NECP2_HUMAN | 34 | 30 | 0.09310 | 1.392 | WT | R10W | Adaptin ear-binding coat-associated protein 2 GN=NECAP2 |
| P36542\|ATPG_HUMAN | 1 | 1 | 0.09356 | 1.869 | R10W | WT | ATP synthase subunit gamma, mitochondrial GN=ATP5F1C |
| P48643\|TCPE_HUMAN | 20 | 19 | 0.09449 | 1.418 | R10W | WT | T-complex protein 1 subunit epsilon GN=CCT5 |
| Q9UBP9\|GULP1_HUMAN | 1 | 1 | 0.09491 | 1.525 | WT | R10W | PTB domain-containing engulfment adapter protein 1 GN=GULP1 |
| P12004\|PCNA_HUMAN | 15 | 15 | 0.09503 | 1.200 | R10W | WT | Proliferating cell nuclear antigen GN=PCNA |
| O76003\|GLRX3_HUMAN | 1 | 1 | 0.09815 | 1.408 | R10W | WT | Glutaredoxin-3 GN=GLRX3 |
| P00736\|C1R_HUMAN | 2 | 2 | 0.09817 | 1.173 | WT | R10W | Complement C1r subcomponent GN=C1R |
| P04406\|G3P_HUMAN | 35 | 33 | 0.09903 | 1.185 | R10W | WT | Glyceraldehyde-3-phosphate dehydrogenase GN=GAPDH |
| Q9Y314\|NOSIP_HUMAN | 1 | 1 | 0.10061 | 1.558 | WT | R10W | Nitric oxide synthase-interacting protein GN=NOSIP |
| P12235\|ADT1_HUMAN | 13 | 1 | 0.10812 | 1.523 | R10W | WT | ADP/ATP translocase 1 GN=SLC25A4 |
| P60842\|IF4A1_HUMAN | 11 | 10 | 0.11124 | 1.359 | R10W | WT | Eukaryotic initiation factor 4A-I GN=EIF4A1 |
| Q53GQ0\|DHB12_HUMAN | 1 | 1 | 0.11565 | 1.422 | R10W | WT | Very-long-chain 3-oxoacyl-CoA reductase GN=HSD17B12 |
| P09496\|CLCA_HUMAN | 1 | 1 | 0.11609 | 1.836 | WT | R10W | Clathrin light chain A GN=CLTA |
| O94973\|AP2A2_HUMAN | 124 | 70 | 0.11783 | 1.327 | WT | R10W | AP-2 complex subunit alpha-2 GN=AP2A2 |
| P62258\|1433E_HUMAN | 8 | 4 | 0.11824 | 1.265 | R10W | WT | 14-3-3 protein epsilon GN=YWHAE |
| P05141\|ADT2_HUMAN | 20 | 3 | 0.12010 | 1.318 | R10W | WT | ADP/ATP translocase 2 GN=SLC25A5 |
| Q15393\|SF3B3_HUMAN | 2 | 2 | 0.12070 | 1.458 | R10W | WT | Splicing factor 3B subunit 3 GN=SF3B3 |
| P50914\|RL14_HUMAN | 1 | 1 | 0.12280 | 1.512 | R10W | WT | 60S ribosomal protein L14 GN=RPL14 |
| Q9UJY1\|HSPB8_HUMAN | 3 | 3 | 0.12533 | 1.277 | R10W | WT | Heat shock protein beta-8 GN=HSPB8 |
| P63122\|VPK8_HUMAN;P63125\|VPK25_HUMAN;P63128\|POK9_HUMAN | 1 | 1 | 0.12627 | 1.138 | R10W | WT | Endogenous retrovirus group K member 8 Pro protein GN=ERVK-8 |
| P60660\|MYL6_HUMAN | 4 | 4 | 0.12736 | 1.631 | R10W | WT | Myosin light polypeptide 6 GN=MYL6 |
| P12956\|XRCC6_HUMAN | 6 | 6 | 0.12749 | 1.359 | R10W | WT | X-ray repair cross-complementing protein 6 GN=XRCC6 |
| P52272\|HNRPM_HUMAN | 7 | 7 | 0.12883 | 1.182 | R10W | WT | Heterogeneous nuclear ribonucleoprotein M GN=HNRNPM |
| O00151\|PDLI1_HUMAN | 1 | 1 | 0.13297 | 1.621 | R10W | WT | PDZ and LIM domain protein 1 GN=PDLIM1 |
| P08559\|ODPA_HUMAN | 1 | 1 | 0.13320 | 1.519 | R10W | WT | Pyruvate dehydrogenase E1 component subunit alpha, somatic form, mitochondrial GN=PDHA1 |
| Q99714\|HCD2_HUMAN | 2 | 2 | 0.14188 | 1.265 | R10W | WT | 3-hydroxyacyl-CoA dehydrogenase type-2 GN=HSD17B10 |
| P27797\|CALR_HUMAN | 1 | 1 | 0.14458 | 1.457 | WT | R10W | Calreticulin GN=CALR |
| Q15084\|PDIA6_HUMAN | 5 | 5 | 0.14740 | 1.282 | R10W | WT | Protein disulfide-isomerase A6 GN=PDIA6 |
| P55084\|ECHB_HUMAN | 1 | 1 | 0.14798 | 1.174 | R10W | WT | Trifunctional enzyme subunit beta, mitochondrial GN=HADHB |
| P53634\|CATC_HUMAN | 1 | 1 | 0.14926 | 1.543 | R10W | WT | Dipeptidyl peptidase 1 GN=CTSC |
| P17844\|DDX5_HUMAN;Q92841\|DDX17_HUMAN | 1 | 1 | 0.15173 | 1.299 | R10W | WT | Probable ATP-dependent RNA helicase DDX5 GN=DDX5 |
| A0A075B6R9\|KVD24_HUMAN;A0A0C4DH68\|KV224_HUMAN | 25 | 16 | 0.15201 | 1.116 | WT | R10W | Probable non-functional immunoglobulin kappa variable 2D-24 GN=IGKV2D-24 PE=5 |
| Q9Y4W6\|AFG32_HUMAN | 2 | 1 | 0.15600 | 1.749 | R10W | WT | AFG3-like protein 2 GN=AFG3L2 |
| P00338\|LDHA_HUMAN | 3 | 2 | 0.15651 | 1.800 | R10W | WT | L-lactate dehydrogenase A chain GN=LDHA |

| | | | | | | | |
|---|---|---|---|---|---|---|---|
| P05109\|S10A8_HUMAN | 1 | 1 | 0.15711 | 4.976 | WT | R10W | Protein S100-A8 GN=S100A8 |
| Q13015\|AF1Q_HUMAN | 1 | 1 | 0.15780 | 1.492 | R10W | WT | Protein AF1q GN=MLLT11 |
| P31689\|DNJA1_HUMAN | 4 | 4 | 0.15844 | 1.409 | R10W | WT | DnaJ homolog subfamily A member 1 GN=DNAJA1 |
| O75616\|ERAL1_HUMAN | 1 | 1 | 0.15934 | 1.302 | WT | R10W | GTPase Era, mitochondrial GN=ERAL1 |
| P62854\|RS26_HUMAN;Q5JNZ5\|RS26L_HUMAN | 2 | 2 | 0.15937 | 1.271 | R10W | WT | 40S ribosomal protein S26 GN=RPS26 |
| Q9UKV5\|AMFR_HUMAN | 1 | 1 | 0.16169 | 2.105 | R10W | WT | E3 ubiquitin-protein ligase AMFR GN=AMFR |
| Q08J23\|NSUN2_HUMAN | 1 | 1 | 0.16341 | 21.114 | R10W | WT | RNA cytosine C(5)-methyltransferase NSUN2 GN=NSUN2 |
| P04083\|ANXA1_HUMAN | 2 | 2 | 0.16421 | 1.821 | R10W | WT | Annexin A1 GN=ANXA1 |
| P06312\|KV401_HUMAN | 27 | 7 | 0.16485 | 1.301 | R10W | WT | Immunoglobulin kappa variable 4-1 GN=IGKV4-1 |
| Q9H853\|TBA4B_HUMAN | 27 | 2 | 0.16533 | 1.725 | R10W | WT | Putative tubulin-like protein alpha-4B GN=TUBA4B PE=5 |
| Q15942\|ZYX_HUMAN | 12 | 12 | 0.16562 | 1.253 | R10W | WT | Zyxin GN=ZYX |
| Q9GZP9\|DERL2_HUMAN | 1 | 1 | 0.16593 | 2.041 | R10W | WT | Derlin-2 GN=DERL2 |
| Q15311\|RBP1_HUMAN | 3 | 3 | 0.17150 | 1.531 | WT | R10W | RalA-binding protein 1 GN=RALBP1 |
| Q3ZCQ8\|TIM50_HUMAN | 15 | 15 | 0.17191 | 1.337 | R10W | WT | Mitochondrial import inner membrane translocase subunit TIM50 GN=TIMM50 |
| P14868\|SYDC_HUMAN | 2 | 2 | 0.17450 | 1.623 | R10W | WT | Aspartate--tRNA ligase, cytoplasmic GN=DARS1 |
| Q86VP6\|CAND1_HUMAN | 1 | 1 | 0.18279 | 1.504 | R10W | WT | Cullin-associated NEDD8-dissociated protein 1 GN=CAND1 |
| Q9Y5X1\|SNX9_HUMAN | 5 | 5 | 0.18352 | 1.429 | WT | R10W | Sorting nexin-9 GN=SNX9 |
| P43686\|PRS6B_HUMAN | 6 | 6 | 0.18536 | 1.284 | R10W | WT | 26S proteasome regulatory subunit 6B GN=PSMC4 |
| P55072\|TERA_HUMAN | 13 | 11 | 0.18550 | 1.873 | R10W | WT | Transitional endoplasmic reticulum ATPase GN=VCP |
| P46782\|RS5_HUMAN | 2 | 1 | 0.18590 | 1.463 | R10W | WT | 40S ribosomal protein S5 GN=RPS5 |
| A0A087WSY4\|HV432_HUMAN | 3 | 1 | 0.19182 | 1.270 | R10W | WT | Immunoglobulin heavy variable 4-30-2 GN=IGHV4-30-2 |
| P07195\|LDHB_HUMAN | 9 | 8 | 0.19383 | 1.173 | WT | R10W | L-lactate dehydrogenase B chain GN=LDHB |
| P39023\|RL3_HUMAN | 5 | 5 | 0.19711 | 1.174 | R10W | WT | 60S ribosomal protein L3 GN=RPL3 |
| Q13596\|SNX1_HUMAN | 1 | 1 | 0.20152 | 1.301 | WT | R10W | Sorting nexin-1 GN=SNX1 |
| Q14192\|FHL2_HUMAN | 2 | 2 | 0.20228 | 1.223 | R10W | WT | Four and a half LIM domains protein 2 GN=FHL2 |
| Q9BWS9\|CHID1_HUMAN | 1 | 1 | 0.20368 | 1.284 | R10W | WT | Chitinase domain-containing protein 1 GN=CHID1 |
| P68371\|TBB4B_HUMAN | 161 | 2 | 0.20518 | 2.015 | R10W | WT | Tubulin beta-4B chain GN=TUBB4B |
| Q06055\|AT5G2_HUMAN | 1 | 1 | 0.20795 | 1.978 | R10W | WT | ATP synthase F(0) complex subunit C2, mitochondrial GN=ATP5MC2 |
| | 60 | 32 | 0.21097 | 1.365 | R10W | WT | |
| P0CG47\|UBB_HUMAN;P0CG48\|UBC_HUMAN;P62979\|RS27A_HUMAN;P62987\|RL40_HUMAN | 1 | 1 | 0.21160 | 1.391 | R10W | WT | Polyubiquitin-B GN=UBB |
| O00743\|PPP6_HUMAN | 3 | 3 | 0.21529 | 1.103 | WT | R10W | Serine/threonine-protein phosphatase 6 catalytic subunit GN=PPP6C |
| Q8NF37\|PCAT1_HUMAN | 1 | 1 | 0.21591 | 1.956 | R10W | WT | Lysophosphatidylcholine acyltransferase 1 GN=LPCAT1 |
| Q8TAT6\|NPL4_HUMAN | 2 | 2 | 0.21787 | 1.573 | R10W | WT | Nuclear protein localization protein 4 homolog GN=NPLOC4 |
| O43684\|BUB3_HUMAN | 1 | 1 | 0.21931 | 1.278 | R10W | WT | Mitotic checkpoint protein BUB3 GN=BUB3 |
| A0A0C4DH26\|KVD41_HUMAN | 11 | 1 | 0.22154 | 1.396 | R10W | WT | Probable non-functional immunoglobulin kappa variable 6D-41 GN=IGKV6D-41 PE=5 |
| Q04323\|UBXN1_HUMAN | 2 | 2 | 0.22288 | 1.281 | R10W | WT | UBX domain-containing protein 1 GN=UBXN1 |
| P35527\|K1C9_HUMAN | 14 | 14 | 0.22925 | 8.057 | WT | R10W | Keratin, type I cytoskeletal 9 GN=KRT9 |
| P53677\|AP3M2_HUMAN | 1 | 1 | 0.23104 | 1.904 | WT | R10W | AP-3 complex subunit mu-2 GN=AP3M2 |
| Q15369\|ELOC_HUMAN | 1 | 1 | 0.23547 | 1.331 | WT | R10W | Elongin-C GN=ELOC |
| Q99470\|SDF2_HUMAN | 2 | 2 | 0.23885 | 1.186 | R10W | WT | Stromal cell-derived factor 2 GN=SDF2 |
| P05783\|K1C18_HUMAN | 8 | 7 | 0.24189 | 1.528 | R10W | WT | Keratin, type I cytoskeletal 18 GN=KRT18 |
| Q53H12\|AGK_HUMAN | 1 | 1 | 0.24524 | 2.215 | R10W | WT | Acylglycerol kinase, mitochondrial GN=AGK |
| P13995\|MTDC_HUMAN | 10 | 10 | 0.24632 | 1.302 | WT | R10W | Bifunctional methylenetetrahydrofolate dehydrogenase/cyclohydrolase, mitochondrial GN=MTHFD2 |
| P05386\|RLA1_HUMAN | 3 | 3 | 0.24748 | 1.197 | R10W | WT | 60S acidic ribosomal protein P1 GN=RPLP1 |
| Q8NC96\|NECP1_HUMAN | 24 | 20 | 0.24765 | 1.361 | WT | R10W | Adaptin ear-binding coat-associated protein 1 GN=NECAP1 |
| P04746\|AMYP_HUMAN | 4 | 4 | 0.24790 | 5.354 | WT | R10W | Pancreatic alpha-amylase GN=AMY2A |
| P33993\|MCM7_HUMAN | 2 | 2 | 0.24835 | 1.279 | R10W | WT | DNA replication licensing factor MCM7 GN=MCM7 |
| P27824\|CALX_HUMAN | 5 | 5 | 0.24892 | 1.285 | R10W | WT | Calnexin GN=CANX |
| Q9BQE3\|TBA1C_HUMAN | 95 | 3 | 0.25920 | 1.115 | R10W | WT | Tubulin alpha-1C chain GN=TUBA1C |
| P68363\|TBA1B_HUMAN | 96 | 1 | 0.25920 | 1.115 | R10W | WT | Tubulin alpha-1B chain GN=TUBA1B |
| Q71U36\|TBA1A_HUMAN | 92 | 2 | 0.25920 | 1.115 | R10W | WT | Tubulin alpha-1A chain GN=TUBA1A |
| P42566\|EPS15_HUMAN | 90 | 88 | 0.25921 | 1.336 | WT | R10W | Epidermal growth factor receptor substrate 15 GN=EPS15 |
| P06576\|ATPB_HUMAN | 14 | 14 | 0.26410 | 1.143 | R10W | WT | ATP synthase subunit beta, mitochondrial GN=ATP5F1B |
| P51170\|SCNNG_HUMAN | 1 | 1 | 0.26842 | 1.258 | R10W | WT | Amiloride-sensitive sodium channel subunit gamma GN=SCNN1G |
| P56545\|CTBP2_HUMAN | 1 | 1 | 0.27213 | 1.185 | R10W | WT | C-terminal-binding protein 2 GN=CTBP2 |
| O14654\|IRS4_HUMAN | 1 | 1 | 0.27860 | 1.676 | WT | R10W | Insulin receptor substrate 4 GN=IRS4 |
| O75880\|SCO1_HUMAN | 1 | 1 | 0.27867 | 1.200 | WT | R10W | Protein SCO1 homolog, mitochondrial GN=SCO1 |
| Q562R1\|ACTBL_HUMAN | 19 | 1 | 0.29179 | 1.316 | R10W | WT | Beta-actin-like protein 2 GN=ACTBL2 |

| | | | | | | | |
|---|---|---|---|---|---|---|---|
| Q9NVI7\|ATD3A_HUMAN | 1 | 1 | 0.29412 | 1.480 | R10W | WT | ATPase family AAA domain-containing protein 3A GN=ATAD3A |
| P22695\|QCR2_HUMAN | 4 | 4 | 0.30082 | 1.404 | R10W | WT | Cytochrome b-c1 complex subunit 2, mitochondrial GN=UQCRC2 |
| P35908\|K22E_HUMAN | 3 | 1 | 0.30499 | 3.162 | WT | R10W | Keratin, type II cytoskeletal 2 epidermal GN=KRT2 |
| P16989\|YBOX3_HUMAN;P67809\|YBOX1_HUMAN | 2 | 2 | 0.30597 | 1.293 | R10W | WT | Y-box-binding protein 3 GN=YBX3 |
| P37802\|TAGL2_HUMAN | 1 | 1 | 0.30932 | 1.426 | R10W | WT | Transgelin-2 GN=TAGLN2 |
| Q8NEZ5\|FBX22_HUMAN | 35 | 34 | 0.31136 | 1.063 | R10W | WT | F-box only protein 22 GN=FBXO22 |
| P55036\|PSMD4_HUMAN | 5 | 5 | 0.31855 | 1.141 | R10W | WT | 26S proteasome non-ATPase regulatory subunit 4 GN=PSMD4 |
| P13804\|ETFA_HUMAN | 1 | 1 | 0.32379 | 1.324 | WT | R10W | Electron transfer flavoprotein subunit alpha, mitochondrial GN=ETFA |
| P29692\|EF1D_HUMAN | 4 | 3 | 0.32613 | 1.230 | R10W | WT | Elongation factor 1-delta GN=EEF1D |
| Q14697\|GANAB_HUMAN | 1 | 1 | 0.32849 | 1.306 | R10W | WT | Neutral alpha-glucosidase AB GN=GANAB |
| P41250\|GARS_HUMAN | 7 | 7 | 0.33295 | 1.161 | WT | R10W | Glycine--tRNA ligase GN=GARS1 |
| P04843\|RPN1_HUMAN | 2 | 2 | 0.33547 | 1.436 | WT | R10W | Dolichyl-diphosphooligosaccharide--protein glycosyltransferase subunit 1 GN=RPN1 |
| A2NJV5\|KV229_HUMAN | 32 | 1 | 0.33699 | 2.918 | R10W | WT | Immunoglobulin kappa variable 2-29 GN=IGKV2-29 |
| O75396\|SC22B_HUMAN | 1 | 1 | 0.34014 | 1.219 | WT | R10W | Vesicle-trafficking protein SEC22b GN=SEC22B |
| P01601\|KVD16_HUMAN | 14 | 2 | 0.34058 | 1.225 | R10W | WT | Immunoglobulin kappa variable 1D-16 GN=IGKV1D-16 |
| P01854\|IGHE_HUMAN | 1 | 1 | 0.34084 | 1.143 | R10W | WT | Immunoglobulin heavy constant epsilon GN=IGHE |
| A0A0C4DH24\|KV621_HUMAN | 13 | 2 | 0.34179 | 1.260 | R10W | WT | Immunoglobulin kappa variable 6-21 GN=IGKV6-21 |
| P23284\|PPIB_HUMAN | 1 | 1 | 0.34624 | 2.122 | R10W | WT | Peptidyl-prolyl cis-trans isomerase B GN=PPIB |
| O75439\|MPPB_HUMAN | 1 | 1 | 0.34782 | 1.909 | WT | R10W | Mitochondrial-processing peptidase subunit beta GN=PMPCB |
| P16520\|GBB3_HUMAN;P62873\|GBB1_HUMAN;P62879\|GBB2_HUMAN | 2 | 2 | 0.34806 | 1.437 | R10W | WT | Guanine nucleotide-binding protein G(I)/G(S)/G(T) subunit beta-3 GN=GNB3 |
| O14561\|ACPM_HUMAN | 1 | 1 | 0.34927 | 1.357 | WT | R10W | Acyl carrier protein, mitochondrial GN=NDUFAB1 |
| P52597\|HNRPF_HUMAN | 4 | 4 | 0.35188 | 1.109 | R10W | WT | Heterogeneous nuclear ribonucleoprotein F GN=HNRNPF |
| Q8WXE9\|STON2_HUMAN | 4 | 4 | 0.35571 | 1.520 | WT | R10W | Stonin-2 GN=STON2 |
| Q00610\|CLH1_HUMAN | 8 | 8 | 0.35579 | 1.173 | R10W | WT | Clathrin heavy chain 1 GN=CLTC |
| P11177\|ODPB_HUMAN | 2 | 2 | 0.36057 | 1.202 | R10W | WT | Pyruvate dehydrogenase E1 component subunit beta, mitochondrial GN=PDHB |
| Q9Y5Y2\|NUBP2_HUMAN | 2 | 2 | 0.36659 | 1.267 | R10W | WT | Cytosolic Fe-S cluster assembly factor NUBP2 GN=NUBP2 |
| Q14694\|UBP10_HUMAN | 1 | 1 | 0.36663 | 1.184 | R10W | WT | Ubiquitin carboxyl-terminal hydrolase 10 GN=USP10 |
| P01859\|IGHG2_HUMAN | 21 | 10 | 0.36718 | 1.422 | R10W | WT | Immunoglobulin heavy constant gamma 2 GN=IGHG2 |
| P06748\|NPM_HUMAN | 7 | 7 | 0.36799 | 1.128 | R10W | WT | Nucleophosmin GN=NPM1 |
| A0A0C4DH34\|HV428_HUMAN | 6 | 3 | 0.36861 | 1.150 | R10W | WT | Immunoglobulin heavy variable 4-28 GN=IGHV4-28 |
| O43143\|DHX15_HUMAN | 1 | 1 | 0.37389 | 1.475 | R10W | WT | Pre-mRNA-splicing factor ATP-dependent RNA helicase DHX15 GN=DHX15 |
| P50225\|ST1A1_HUMAN | 2 | 2 | 0.37626 | 1.241 | WT | R10W | Sulfotransferase 1A1 GN=SULT1A1 |
| Q13347\|EIF3I_HUMAN | 2 | 2 | 0.37710 | 1.152 | R10W | WT | Eukaryotic translation initiation factor 3 subunit I GN=EIF3I |
| P05387\|RLA2_HUMAN | 3 | 3 | 0.37856 | 1.132 | R10W | WT | 60S acidic ribosomal protein P2 GN=RPLP2 |
| P08708\|RS17_HUMAN | 3 | 3 | 0.37868 | 1.111 | R10W | WT | 40S ribosomal protein S17 GN=RPS17 |
| Q9NR30\|DDX21_HUMAN | 2 | 2 | 0.37918 | 1.210 | R10W | WT | Nucleolar RNA helicase 2 GN=DDX21 |
| Q07021\|C1QBP_HUMAN | 1 | 1 | 0.38088 | 1.343 | R10W | WT | Complement component 1 Q subcomponent-binding protein, mitochondrial GN=C1QBP |
| P23246\|SFPQ_HUMAN | 3 | 2 | 0.38167 | 1.061 | R10W | WT | Splicing factor, proline- and glutamine-rich GN=SFPQ |
| P43490\|NAMPT_HUMAN | 1 | 1 | 0.38377 | 1.475 | R10W | WT | Nicotinamide phosphoribosyltransferase GN=NAMPT |
| P41091\|IF2G_HUMAN;Q2VIR3\|IF2GL_HUMAN | 1 | 1 | 0.38406 | 1.450 | WT | R10W | Eukaryotic translation initiation factor 2 subunit 3 GN=EIF2S3 |
| Q658Y4\|F91A1_HUMAN | 1 | 1 | 0.38481 | 1.229 | R10W | WT | Protein FAM91A1 GN=FAM91A1 |
| P04433\|KV311_HUMAN | 12 | 2 | 0.38511 | 1.173 | R10W | WT | Immunoglobulin kappa variable 3-11 GN=IGKV3-11 |
| Q10713\|MPPA_HUMAN | 1 | 1 | 0.38573 | 1.413 | WT | R10W | Mitochondrial-processing peptidase subunit alpha GN=PMPCA |
| P13010\|XRCC5_HUMAN | 4 | 4 | 0.38810 | 1.271 | WT | R10W | X-ray repair cross-complementing protein 5 GN=XRCC5 |
| P11586\|C1TC_HUMAN | 1 | 1 | 0.39202 | 1.346 | WT | R10W | C-1-tetrahydrofolate synthase, cytoplasmic GN=MTHFD1 |
| Q9BSD7\|NTPCR_HUMAN | 4 | 4 | 0.39547 | 1.142 | R10W | WT | Cancer-related nucleoside-triphosphatase GN=NTPCR |
| P68104\|EF1A1_HUMAN | 79 | 79 | 0.39584 | 1.181 | R10W | WT | Elongation factor 1-alpha 1 GN=EEF1A1 |
| P62701\|RS4X_HUMAN | 8 | 8 | 0.39787 | 1.478 | R10W | WT | 40S ribosomal protein S4, X isoform GN=RPS4X |
| P58876\|H2B1D_HUMAN;P62807\|H2B1C_HUMAN;Q5QNW6\|H2B2F_HUMAN;Q93079\|H2B1H_HUMAN;Q99877\|H2B1N_HUMAN;Q99879\|H2B1M_HUMAN | 21 | 21 | 0.39815 | 1.268 | R10W | WT | Histone H2B type 1-D GN=H2BC5 |
| Q02790\|FKBP4_HUMAN | 2 | 2 | 0.39916 | 1.208 | R10W | WT | Peptidyl-prolyl cis-trans isomerase FKBP4 GN=FKBP4 |
| O95373\|IPO7_HUMAN | 1 | 1 | 0.39973 | 1.433 | R10W | WT | Importin-7 GN=IPO7 |
| P17987\|TCPA_HUMAN | 7 | 6 | 0.40208 | 1.213 | R10W | WT | T-complex protein 1 subunit alpha GN=TCP1 |
| Q14204\|DYHC1_HUMAN | 1 | 1 | 0.40464 | 1.218 | R10W | WT | Cytoplasmic dynein 1 heavy chain 1 GN=DYNC1H1 |
| Q9UBB4\|ATX10_HUMAN | 1 | 1 | 0.40557 | 1.138 | R10W | WT | Ataxin-10 GN=ATXN10 |
| P09936\|UCHL1_HUMAN | 2 | 2 | 0.40937 | 1.060 | R10W | WT | Ubiquitin carboxyl-terminal hydrolase isozyme L1 GN=UCHL1 |
| P08670\|VIME_HUMAN | 3 | 2 | 0.40978 | 1.298 | R10W | WT | Vimentin GN=VIM |
| Q7L1W4\|LRC8D_HUMAN | 1 | 1 | 0.41177 | 1.169 | R10W | WT | Volume-regulated anion channel subunit LRRC8D GN=LRRC8D |

| | | | | | | | |
|---|---|---|---|---|---|---|---|
| P56945\|BCAR1_HUMAN | 52 | 50 | 0.42314 | 1.378 | R10W | WT | Breast cancer anti-estrogen resistance protein 1 GN=BCAR1 |
| A0A075B6I0\|LV861_HUMAN | 1 | 1 | 0.42517 | 1.206 | R10W | WT | Immunoglobulin lambda variable 8-61 GN=IGLV8-61 |
| Q5H9R7\|PP6R3_HUMAN | 3 | 3 | 0.42589 | 1.480 | WT | R10W | Serine/threonine-protein phosphatase 6 regulatory subunit 3 GN=PPP6R3 |
| P0C0S8\|H2A1_HUMAN;P20671\|H2A1D_HUMAN;Q16777\|H2A2C_HUMAN;Q6FI13\|H2A2A_HUMAN;Q96KK5\|H2A1H_HUMAN;Q99878\|H2A1J_HUMAN;Q9BTM1\|H2AJ_HUMAN | 9 | 8 | 0.42674 | 1.479 | R10W | WT | Histone H2A type 1 GN=H2AC11 |
| P14618\|KPYM_HUMAN | 53 | 52 | 0.42712 | 1.163 | WT | R10W | Pyruvate kinase PKM GN=PKM |
| Q9HCC0\|MCCB_HUMAN | 2 | 2 | 0.42713 | 1.154 | WT | R10W | Methylcrotonoyl-CoA carboxylase beta chain, mitochondrial GN=MCCC2 |
| Q15366\|PCBP2_HUMAN | 11 | 6 | 0.42765 | 1.150 | R10W | WT | Poly(rC)-binding protein 2 GN=PCBP2 |
| Q13011\|ECH1_HUMAN | 5 | 5 | 0.43046 | 1.205 | WT | R10W | Delta(3,5)-Delta(2,4)-dienoyl-CoA isomerase, mitochondrial GN=ECH1 |
| P25705\|ATPA_HUMAN | 23 | 23 | 0.43273 | 1.249 | R10W | WT | ATP synthase subunit alpha, mitochondrial GN=ATP5F1A |
| P23378\|GCSP_HUMAN | 1 | 1 | 0.43495 | 1.314 | R10W | WT | Glycine dehydrogenase (decarboxylating), mitochondrial GN=GLDC |
| Q2M2I8\|AAK1_HUMAN | 12 | 10 | 0.43658 | 1.302 | WT | R10W | AP2-associated protein kinase 1 GN=AAK1 |
| P68871\|HBB_HUMAN | 8 | 8 | 0.44261 | 1.270 | R10W | WT | Hemoglobin subunit beta GN=HBB |
| Q15233\|NONO_HUMAN | 7 | 6 | 0.44423 | 1.198 | R10W | WT | Non-POU domain-containing octamer-binding protein GN=NONO |
| P50395\|GDIB_HUMAN | 1 | 1 | 0.44494 | 1.288 | R10W | WT | Rab GDP dissociation inhibitor beta GN=GDI2 |
| P18124\|RL7_HUMAN | 4 | 3 | 0.44531 | 1.202 | R10W | WT | 60S ribosomal protein L7 GN=RPL7 |
| P15924\|DESP_HUMAN | 3 | 3 | 0.45113 | 1.781 | R10W | WT | Desmoplakin GN=DSP |
| Q9Y2I6\|NINL_HUMAN | 1 | 1 | 0.45515 | 1.192 | WT | R10W | Ninein-like protein GN=NINL |
| P62805\|H4_HUMAN | 4 | 4 | 0.45584 | 1.394 | R10W | WT | Histone H4 GN=H4C1 |
| P09651\|ROA1_HUMAN;Q32P51\|RA1L2_HUMAN | 1 | 1 | 0.46235 | 1.156 | R10W | WT | Heterogeneous nuclear ribonucleoprotein A1 GN=HNRNPA1 |
| P62753\|RS6_HUMAN | 3 | 3 | 0.46947 | 1.165 | WT | R10W | 40S ribosomal protein S6 GN=RPS6 |
| P03254\|E1A_ADE02;P03255\|E1A_ADE05 | 1 | 1 | 0.47068 | 2.247 | WT | R10W | Early E1A protein OS=Human adenovirus C serotype 2 OX=10515 |
| P51571\|SSRD_HUMAN | 2 | 2 | 0.47211 | 1.132 | R10W | WT | Translocon-associated protein subunit delta GN=SSR4 |
| Q15758\|AAAT_HUMAN | 1 | 1 | 0.47245 | 1.168 | R10W | WT | Neutral amino acid transporter B(0) GN=SLC1A5 |
| P19623\|SPEE_HUMAN | 4 | 4 | 0.47427 | 1.163 | R10W | WT | Spermidine synthase GN=SRM |
| O75531\|BAF_HUMAN | 1 | 1 | 0.47745 | 4.781 | WT | R10W | Barrier-to-autointegration factor GN=BANF1 |
| P07355\|ANXA2_HUMAN | 3 | 3 | 0.48178 | 1.666 | R10W | WT | Annexin A2 GN=ANXA2 |
| O00244\|ATOX1_HUMAN | 1 | 1 | 0.48293 | 1.147 | WT | R10W | Copper transport protein ATOX1 GN=ATOX1 |
| Q494V2\|CP100_HUMAN | 1 | 1 | 0.48715 | 1.565 | R10W | WT | Cilia- and flagella-associated protein 100 GN=CFAP100 |
| O95678\|K2C75_HUMAN | 4 | 1 | 0.48818 | 1.105 | R10W | WT | Keratin, type II cytoskeletal 75 GN=KRT75 |
| O00370\|LORF2_HUMAN | 1 | 1 | 0.49183 | 1.151 | R10W | WT | LINE-1 retrotransposable element ORF2 protein |
| Q13643\|FHL3_HUMAN | 3 | 3 | 0.49274 | 1.250 | R10W | WT | Four and a half LIM domains protein 3 GN=FHL3 |
| P22061\|PIMT_HUMAN | 25 | 25 | 0.49299 | 1.095 | R10W | WT | Protein-L-isoaspartate(D-aspartate) O-methyltransferase GN=PCMT1 |
| P05388\|RLA0_HUMAN | 3 | 3 | 0.49387 | 1.262 | R10W | WT | 60S acidic ribosomal protein P0 GN=RPLP0 |
| P62266\|RS23_HUMAN | 1 | 1 | 0.49405 | 1.243 | WT | R10W | 40S ribosomal protein S23 GN=RPS23 |
| P07437\|TBB5_HUMAN | 163 | 10 | 0.50719 | 1.077 | R10W | WT | Tubulin beta chain GN=TUBB |
| Q06210\|GFPT1_HUMAN | 1 | 1 | 0.50973 | 1.083 | R10W | WT | Glutamine--fructose-6-phosphate aminotransferase [isomerizing] 1 GN=GFPT1 |
| Q16576\|RBBP7_HUMAN | 7 | 7 | 0.51098 | 1.145 | WT | R10W | Histone-binding protein RBBP7 GN=RBBP7 |
| Q9BRK5\|CAB45_HUMAN | 2 | 2 | 0.51506 | 1.182 | R10W | WT | 45 kDa calcium-binding protein GN=SDF4 |
| P50991\|TCPD_HUMAN | 9 | 8 | 0.51627 | 1.161 | R10W | WT | T-complex protein 1 subunit delta GN=CCT4 |
| Q99832\|TCPH_HUMAN | 7 | 6 | 0.52128 | 1.240 | R10W | WT | T-complex protein 1 subunit eta GN=CCT7 |
| Q9Y305\|ACOT9_HUMAN | 7 | 7 | 0.52466 | 1.133 | WT | R10W | Acyl-coenzyme A thioesterase 9, mitochondrial GN=ACOT9 |
| P69905\|HBA_HUMAN | 7 | 7 | 0.52486 | 1.381 | R10W | WT | Hemoglobin subunit alpha GN=HBA1 |
| Q9P258\|RCC2_HUMAN | 5 | 5 | 0.52635 | 1.045 | WT | R10W | Protein RCC2 GN=RCC2 |
| Q9UM54\|MYO6_HUMAN | 1 | 1 | 0.53250 | 1.346 | R10W | WT | Unconventional myosin-VI GN=MYO6 |
| Q14207\|NPAT_HUMAN | 1 | 1 | 0.53308 | 1.175 | R10W | WT | Protein NPAT GN=NPAT |
| P62906\|RL10A_HUMAN | 6 | 6 | 0.53357 | 1.145 | R10W | WT | 60S ribosomal protein L10a GN=RPL10A |
| Q96CW1\|AP2M1_HUMAN | 95 | 92 | 0.53416 | 1.333 | WT | R10W | AP-2 complex subunit mu GN=AP2M1 |
| Q9Y266\|NUDC_HUMAN | 2 | 2 | 0.53566 | 1.117 | R10W | WT | Nuclear migration protein nudC GN=NUDC |
| P53597\|SUCA_HUMAN | 1 | 1 | 0.54125 | 1.266 | WT | R10W | Succinate--CoA ligase [ADP/GDP-forming] subunit alpha, mitochondrial GN=SUCLG1 |
| Q9BRP1\|PDD2L_HUMAN | 1 | 1 | 0.54149 | 1.125 | R10W | WT | Programmed cell death protein 2-like GN=PDCD2L |
| P54136\|SYRC_HUMAN | 1 | 1 | 0.54639 | 1.151 | R10W | WT | Arginine--tRNA ligase, cytoplasmic GN=RARS1 |
| P60709\|ACTB_HUMAN;P63261\|ACTG_HUMAN | 40 | 20 | 0.54897 | 1.103 | R10W | WT | Actin, cytoplasmic 1 GN=ACTB |
| Q07065\|CKAP4_HUMAN | 7 | 7 | 0.54924 | 1.137 | WT | R10W | Cytoskeleton-associated protein 4 GN=CKAP4 |
| P07737\|PROF1_HUMAN | 1 | 1 | 0.54940 | 1.558 | R10W | WT | Profilin-1 GN=PFN1 |
| O14818\|PSA7_HUMAN | 1 | 1 | 0.54941 | 1.117 | R10W | WT | Proteasome subunit alpha type-7 GN=PSMA7 |
| P35579\|MYH9_HUMAN;P35580\|MYH10_HUMAN | 1 | 1 | 0.55104 | 1.082 | R10W | WT | Myosin-9 GN=MYH9 |
| P10809\|CH60_HUMAN | 71 | 71 | 0.55689 | 1.235 | R10W | WT | 60 kDa heat shock protein, mitochondrial GN=HSPD1 |
| Q96JB1\|DYH8_HUMAN | 1 | 1 | 0.55739 | 1.138 | WT | R10W | Dynein heavy chain 8, axonemal GN=DNAH8 |

| | | | | | | | |
|---|---|---|---|---|---|---|---|
| A0A075B6S6\|KVD30_HUMAN;P06310\|KV230_HUMAN | 52 | 21 | 0.56094 | 1.244 | R10W | WT | Immunoglobulin kappa variable 2D-30 GN=IGKV2D-30 |
| P35998\|PRS7_HUMAN | 4 | 4 | 0.56210 | 1.094 | R10W | WT | 26S proteasome regulatory subunit 7 GN=PSMC2 |
| Q9BUF5\|TBB6_HUMAN | 85 | 5 | 0.56372 | 1.334 | R10W | WT | Tubulin beta-6 chain GN=TUBB6 |
| O43426\|SYNJ1_HUMAN | 3 | 3 | 0.56428 | 1.164 | WT | R10W | Synaptojanin-1 GN=SYNJ1 |
| P0DMV8\|HS71A_HUMAN;P0DMV9\|HS71B_HUMAN | 121 | 55 | 0.56464 | 1.103 | WT | R10W | Heat shock 70 kDa protein 1A GN=HSPA1A |
| P26038\|MOES_HUMAN | 1 | 1 | 0.56762 | 1.137 | WT | R10W | Moesin GN=MSN |
| P27635\|RL10_HUMAN | 2 | 2 | 0.57192 | 1.256 | R10W | WT | 60S ribosomal protein L10 GN=RPL10 |
| P50454\|SERPH_HUMAN | 9 | 8 | 0.57503 | 1.257 | R10W | WT | Serpin H1 GN=SERPINH1 |
| P34897\|GLYM_HUMAN | 2 | 2 | 0.57518 | 1.184 | WT | R10W | Serine hydroxymethyltransferase, mitochondrial GN=SHMT2 |
| P08865\|RSSA_HUMAN | 2 | 2 | 0.57804 | 1.240 | R10W | WT | 40S ribosomal protein SA GN=RPSA |
| Q13616\|CUL1_HUMAN | 1 | 1 | 0.57805 | 1.334 | R10W | WT | Cullin-1 GN=CUL1 |
| P04181\|OAT_HUMAN | 12 | 12 | 0.58085 | 1.147 | WT | R10W | Ornithine aminotransferase, mitochondrial GN=OAT |
| Q15370\|ELOB_HUMAN | 4 | 4 | 0.58674 | 1.208 | R10W | WT | Elongin-B GN=ELOB |
| P36578\|RL4_HUMAN | 1 | 1 | 0.59200 | 1.162 | R10W | WT | 60S ribosomal protein L4 GN=RPL4 |
| Q06830\|PRDX1_HUMAN | 10 | 10 | 0.59286 | 1.055 | WT | R10W | Peroxiredoxin-1 GN=PRDX1 |
| P62318\|SMD3_HUMAN | 1 | 1 | 0.59468 | 1.515 | R10W | WT | Small nuclear ribonucleoprotein Sm D3 GN=SNRPD3 |
| P62910\|RL32_HUMAN | 1 | 1 | 0.59608 | 1.121 | WT | R10W | 60S ribosomal protein L32 GN=RPL32 |
| Q6PCT2\|FXL19_HUMAN | 1 | 1 | 0.59718 | 1.130 | WT | R10W | F-box/LRR-repeat protein 19 GN=FBXL19 |
| Q13885\|TBB2A_HUMAN | 140 | 4 | 0.59888 | 1.237 | R10W | WT | Tubulin beta-2A chain GN=TUBB2A |
| P51114\|FXR1_HUMAN | 1 | 1 | 0.60230 | 1.105 | WT | R10W | Fragile X mental retardation syndrome-related protein 1 GN=FXR1 |
| P30050\|RL12_HUMAN | 4 | 4 | 0.61032 | 1.091 | R10W | WT | 60S ribosomal protein L12 GN=RPL12 |
| P61978\|HNRPK_HUMAN | 8 | 8 | 0.61410 | 1.108 | R10W | WT | Heterogeneous nuclear ribonucleoprotein K GN=HNRNPK |
| P46778\|RL21_HUMAN | 2 | 2 | 0.61794 | 1.090 | R10W | WT | 60S ribosomal protein L21 GN=RPL21 |
| O95817\|BAG3_HUMAN | 1 | 1 | 0.61902 | 1.174 | R10W | WT | BAG family molecular chaperone regulator 3 GN=BAG3 |
| P78371\|TCPB_HUMAN | 10 | 8 | 0.62479 | 1.056 | WT | R10W | T-complex protein 1 subunit beta GN=CCT2 |
| P62249\|RS16_HUMAN | 2 | 2 | 0.62628 | 1.104 | R10W | WT | 40S ribosomal protein S16 GN=RPS16 |
| P0DP23\|CALM1_HUMAN;P0DP24\|CALM2_HUMAN;P0DP25\|CALM3_HUMAN | 4 | 4 | 0.62661 | 1.089 | R10W | WT | Calmodulin-1 GN=CALM1 |
| Q8ND24\|RN214_HUMAN | 1 | 1 | 0.62699 | 1.155 | R10W | WT | RING finger protein 214 GN=RNF214 |
| P14174\|MIF_HUMAN | 3 | 3 | 0.62721 | 1.049 | WT | R10W | Macrophage migration inhibitory factor GN=MIF |
| P12277\|KCRB_HUMAN | 6 | 6 | 0.62875 | 1.088 | R10W | WT | Creatine kinase B-type GN=CKB |
| P42704\|LPPRC_HUMAN | 1 | 1 | 0.62912 | 1.258 | WT | R10W | Leucine-rich PPR motif-containing protein, mitochondrial GN=LRPPRC |
| Q12962\|TAF10_HUMAN | 1 | 1 | 0.62927 | 1.051 | R10W | WT | Transcription initiation factor TFIID subunit 10 GN=TAF10 |
| Q14576\|ELAV3_HUMAN | 1 | 1 | 0.63355 | 1.245 | R10W | WT | ELAV-like protein 3 GN=ELAVL3 |
| P62899\|RL31_HUMAN | 1 | 1 | 0.64659 | 1.054 | R10W | WT | 60S ribosomal protein L31 GN=RPL31 |
| P11908\|PRPS2_HUMAN;P60891\|PRPS1_HUMAN | 3 | 3 | 0.65012 | 1.063 | R10W | WT | Ribose-phosphate pyrophosphokinase 2 GN=PRPS2 |
| P07814\|SYEP_HUMAN | 1 | 1 | 0.65218 | 1.068 | R10W | WT | Bifunctional glutamate/proline--tRNA ligase GN=EPRS1 |
| P52701\|MSH6_HUMAN | 1 | 1 | 0.65316 | 1.223 | R10W | WT | DNA mismatch repair protein Msh6 GN=MSH6 |
| P49207\|RL34_HUMAN | 2 | 2 | 0.65398 | 1.100 | WT | R10W | 60S ribosomal protein L34 GN=RPL34 |
| P02679\|FIBG_HUMAN | 4 | 4 | 0.66108 | 1.182 | R10W | WT | Fibrinogen gamma chain GN=FGG |
| P62269\|RS18_HUMAN | 1 | 1 | 0.66392 | 1.080 | R10W | WT | 40S ribosomal protein S18 GN=RPS18 |
| P07858\|CATB_HUMAN | 1 | 1 | 0.66442 | 2.121 | WT | R10W | Cathepsin B GN=CTSB |
| P50213\|IDH3A_HUMAN | 5 | 5 | 0.66656 | 1.053 | WT | R10W | Isocitrate dehydrogenase [NAD] subunit alpha, mitochondrial GN=IDH3A |
| P11021\|BIP_HUMAN | 13 | 8 | 0.66726 | 1.070 | R10W | WT | Endoplasmic reticulum chaperone BiP GN=HSPA5 |
| P07686\|HEXB_HUMAN | 1 | 1 | 0.66769 | 1.194 | R10W | WT | Beta-hexosaminidase subunit beta GN=HEXB |
| P14209\|CD99_HUMAN | 1 | 1 | 0.66802 | 1.257 | WT | R10W | CD99 antigen GN=CD99 |
| P49915\|GUAA_HUMAN | 1 | 1 | 0.67824 | 1.356 | R10W | WT | GMP synthase [glutamine-hydrolyzing] GN=GMPS |
| P56192\|SYMC_HUMAN | 1 | 1 | 0.68349 | 1.420 | R10W | WT | Methionine--tRNA ligase, cytoplasmic GN=MARS1 |
| P32969\|RL9_HUMAN | 4 | 4 | 0.68413 | 1.043 | R10W | WT | 60S ribosomal protein L9 GN=RPL9 |
| P38646\|GRP75_HUMAN | 26 | 15 | 0.68706 | 1.031 | R10W | WT | Stress-70 protein, mitochondrial GN=HSPA9 |
| P62937\|PPIA_HUMAN | 18 | 17 | 0.69534 | 1.061 | R10W | WT | Peptidyl-prolyl cis-trans isomerase A GN=PPIA |
| O75400\|PR40A_HUMAN | 1 | 1 | 0.69629 | 1.910 | WT | R10W | Pre-mRNA-processing factor 40 homolog A GN=PRPF40A |
| P22314\|UBA1_HUMAN | 3 | 3 | 0.69735 | 1.118 | WT | R10W | Ubiquitin-like modifier-activating enzyme 1 GN=UBA1 |
| Q6ZSR9\|YJ005_HUMAN | 6 | 6 | 0.71085 | 1.076 | WT | R10W | Uncharacterized protein FLJ45252 |
| P34932\|HSP74_HUMAN | 3 | 3 | 0.71163 | 1.075 | R10W | WT | Heat shock 70 kDa protein 4 GN=HSPA4 |
| O43175\|SERA_HUMAN | 6 | 6 | 0.71407 | 1.077 | WT | R10W | D-3-phosphoglycerate dehydrogenase GN=PHGDH |
| P62424\|RL7A_HUMAN | 5 | 5 | 0.71452 | 1.049 | R10W | WT | 60S ribosomal protein L7a GN=RPL7A |
| Q14974\|IMB1_HUMAN | 7 | 7 | 0.71457 | 1.154 | R10W | WT | Importin subunit beta-1 GN=KPNB1 |
| P78527\|PRKDC_HUMAN | 8 | 8 | 0.71711 | 1.042 | R10W | WT | DNA-dependent protein kinase catalytic subunit GN=PRKDC |
| P26583\|HMGB2_HUMAN | 1 | 1 | 0.72265 | 1.066 | R10W | WT | High mobility group protein B2 GN=HMGB2 |

| | | | | | | | |
|---|---|---|---|---|---|---|---|
| P49458\|SRP09_HUMAN | 1 | 1 | 0.72480 | 1.007 | R10W | WT | Signal recognition particle 9 kDa protein GN=SRP9 |
| Q92854\|SEM4D_HUMAN | 1 | 1 | 0.72534 | 1.325 | R10W | WT | Semaphorin-4D GN=SEMA4D |
| P32119\|PRDX2_HUMAN | 1 | 1 | 0.72552 | 1.172 | R10W | WT | Peroxiredoxin-2 GN=PRDX2 |
| P01715\|LV301_HUMAN | 4 | 2 | 0.72632 | 1.240 | R10W | WT | Immunoglobulin lambda variable 3-1 GN=IGLV3-1 |
| Q15365\|PCBP1_HUMAN | 13 | 8 | 0.72778 | 1.110 | R10W | WT | Poly(rC)-binding protein 1 GN=PCBP1 |
| O15371\|EIF3D_HUMAN | 1 | 1 | 0.73203 | 1.018 | R10W | WT | Eukaryotic translation initiation factor 3 subunit D GN=EIF3D |
| A0A075B6S2\|KVD29_HUMAN | 33 | 2 | 0.73447 | 1.186 | R10W | WT | Immunoglobulin kappa variable 2D-29 GN=IGKV2D-29 |
| A8MWP6\|YQ019_HUMAN;A8MXK9\|YQ018_HUMAN | 1 | 1 | 0.73718 | 1.063 | R10W | WT | Uncharacterized protein ENSP00000382042 |
| O43776\|SYNC_HUMAN | 1 | 1 | 0.73830 | 1.048 | WT | R10W | Asparagine--tRNA ligase, cytoplasmic GN=NARS1 |
| P41180\|CASR_HUMAN | 4 | 4 | 0.74118 | 1.072 | R10W | WT | Extracellular calcium-sensing receptor GN=CASR |
| Q9UKN7\|MYO15_HUMAN | 1 | 1 | 0.74172 | 1.733 | WT | R10W | Unconventional myosin-XV GN=MYO15A |
| E9PAV3\|NACAM_HUMAN;Q13765\|NACA_HUMAN | 1 | 1 | 0.74465 | 1.051 | WT | R10W | Nascent polypeptide-associated complex subunit alpha, muscle-specific form GN=NACA |
| P29120\|NEC1_HUMAN | 1 | 1 | 0.74573 | 1.044 | R10W | WT | Neuroendocrine convertase 1 GN=PCSK1 |
| A0A0B4J1U7\|HV601_HUMAN | 3 | 3 | 0.74617 | 1.062 | R10W | WT | Immunoglobulin heavy variable 6-1 GN=IGHV6-1 |
| P51570\|GALK1_HUMAN | 5 | 5 | 0.74721 | 1.150 | R10W | WT | Galactokinase GN=GALK1 |
| Q00839\|HNRPU_HUMAN | 2 | 2 | 0.74827 | 1.249 | R10W | WT | Heterogeneous nuclear ribonucleoprotein U GN=HNRNPU |
| P31040\|SDHA_HUMAN | 2 | 2 | 0.75101 | 1.067 | WT | R10W | Succinate dehydrogenase [ubiquinone] flavoprotein subunit, mitochondrial GN=SDHA |
| P26599\|PTBP1_HUMAN | 2 | 2 | 0.75210 | 1.083 | R10W | WT | Polypyrimidine tract-binding protein 1 GN=PTBP1 |
| P43487\|RANG_HUMAN | 6 | 6 | 0.75286 | 1.100 | R10W | WT | Ran-specific GTPase-activating protein GN=RANBP1 |
| O60884\|DNJA2_HUMAN | 1 | 1 | 0.75328 | 1.154 | R10W | WT | DnaJ homolog subfamily A member 2 GN=DNAJA2 |
| Q99550\|MPP9_HUMAN | 1 | 1 | 0.75396 | 1.617 | R10W | WT | M-phase phosphoprotein 9 GN=MPHOSPH9 |
| P01824\|HV439_HUMAN | 4 | 1 | 0.76077 | 1.080 | R10W | WT | Immunoglobulin heavy variable 4-39 GN=IGHV4-39 |
| O43852\|CALU_HUMAN | 1 | 1 | 0.76119 | 1.067 | WT | R10W | Calumenin GN=CALU |
| Q149M9\|NWD1_HUMAN | 1 | 1 | 0.76231 | 1.029 | R10W | WT | NACHT domain- and WD repeat-containing protein 1 GN=NWD1 |
| P23528\|COF1_HUMAN | 6 | 6 | 0.76276 | 1.035 | R10W | WT | Cofilin-1 GN=CFL1 |
| Q99426\|TBCB_HUMAN | 2 | 2 | 0.76411 | 1.175 | WT | R10W | Tubulin-folding cofactor B GN=TBCB |
| Q8NC51\|PAIRB_HUMAN | 1 | 1 | 0.77397 | 1.150 | R10W | WT | Plasminogen activator inhibitor 1 RNA-binding protein GN=SERBP1 |
| P63173\|RL38_HUMAN | 2 | 2 | 0.77871 | 1.128 | WT | R10W | 60S ribosomal protein L38 GN=RPL38 |
| O00483\|NDUA4_HUMAN | 2 | 2 | 0.78232 | 1.895 | R10W | WT | Cytochrome c oxidase subunit NDUFA4 GN=NDUFA4 |
| Q14195\|DPYL3_HUMAN | 2 | 2 | 0.78537 | 1.072 | WT | R10W | Dihydropyrimidinase-related protein 3 GN=DPYSL3 |
| Q10587\|TEF_HUMAN | 1 | 1 | 0.78551 | 1.069 | WT | R10W | Thyrotroph embryonic factor GN=TEF |
| P30837\|AL1B1_HUMAN | 1 | 1 | 0.78604 | 1.133 | WT | R10W | Aldehyde dehydrogenase X, mitochondrial GN=ALDH1B1 |
| P07900\|HS90A_HUMAN | 23 | 8 | 0.78794 | 1.125 | WT | R10W | Heat shock protein HSP 90-alpha GN=HSP90AA1 |
| O75306\|NDUS2_HUMAN | 2 | 2 | 0.78910 | 1.075 | R10W | WT | NADH dehydrogenase [ubiquinone] iron-sulfur protein 2, mitochondrial GN=NDUFS2 |
| P61247\|RS3A_HUMAN | 1 | 1 | 0.78998 | 1.036 | R10W | WT | 40S ribosomal protein S3a GN=RPS3A |
| O60547\|GMDS_HUMAN | 4 | 4 | 0.79115 | 1.056 | R10W | WT | GDP-mannose 4,6 dehydratase GN=GMDS |
| P49327\|FAS_HUMAN | 4 | 4 | 0.79134 | 1.242 | R10W | WT | Fatty acid synthase GN=FASN |
| O95831\|AIFM1_HUMAN | 1 | 1 | 0.79816 | 1.051 | R10W | WT | Apoptosis-inducing factor 1, mitochondrial GN=AIFM1 |
| P27348\|1433T_HUMAN | 4 | 1 | 0.80335 | 1.041 | R10W | WT | 14-3-3 protein theta GN=YWHAQ |
| Q9Y285\|SYFA_HUMAN | 2 | 2 | 0.80522 | 1.117 | R10W | WT | Phenylalanine--tRNA ligase alpha subunit GN=FARSA |
| P22087\|FBRL_HUMAN | 1 | 1 | 0.80730 | 1.071 | R10W | WT | rRNA 2'-O-methyltransferase fibrillarin GN=FBL |
| Q96AB3\|ISOC2_HUMAN | 1 | 1 | 0.81398 | 1.028 | WT | R10W | Isochorismatase domain-containing protein 2 GN=ISOC2 |
| P08238\|HS90B_HUMAN | 39 | 22 | 0.81561 | 1.055 | R10W | WT | Heat shock protein HSP 90-beta GN=HSP90AB1 |
| P30101\|PDIA3_HUMAN | 1 | 1 | 0.81579 | 1.095 | WT | R10W | Protein disulfide-isomerase A3 GN=PDIA3 |
| P52339\|DNBI_HHV7J | 1 | 1 | 0.81798 | 1.364 | R10W | WT | Major DNA-binding protein OS=Human herpesvirus 7 (strain JI) OX=57278 GN=DBP |
| P00367\|DHE3_HUMAN | 4 | 4 | 0.81800 | 1.075 | R10W | WT | Glutamate dehydrogenase 1, mitochondrial GN=GLUD1 |
| P30048\|PRDX3_HUMAN | 6 | 6 | 0.82078 | 1.108 | R10W | WT | Thioredoxin-dependent peroxide reductase, mitochondrial GN=PRDX3 |
| P42167\|LAP2B_HUMAN | 1 | 1 | 0.82231 | 1.050 | WT | R10W | Lamina-associated polypeptide 2, isoforms beta/gamma GN=TMPO |
| O14734\|ACOT8_HUMAN | 2 | 2 | 0.82451 | 1.390 | R10W | WT | Acyl-coenzyme A thioesterase 8 GN=ACOT8 |
| P62841\|RS15_HUMAN | 4 | 4 | 0.82631 | 1.083 | WT | R10W | 40S ribosomal protein S15 GN=RPS15 |
| P62241\|RS8_HUMAN | 4 | 4 | 0.82748 | 1.014 | WT | R10W | 40S ribosomal protein S8 GN=RPS8 |
| Q5XKP0\|MIC13_HUMAN | 2 | 1 | 0.83017 | 1.046 | R10W | WT | MICOS complex subunit MIC13 GN=MICOS13 |
| P02671\|FIBA_HUMAN | 2 | 1 | 0.83823 | 1.077 | R10W | WT | Fibrinogen alpha chain GN=FGA |
| P01768\|HV330_HUMAN;P0DP02\|HVC33_HUMAN;P0DP03\|HVC05_HUMAN | 4 | 1 | 0.84069 | 1.094 | R10W | WT | Immunoglobulin heavy variable 3-30 GN=IGHV3-30 |
| P02786\|TFR1_HUMAN | 1 | 1 | 0.84190 | 1.205 | R10W | WT | Transferrin receptor protein 1 GN=TFRC |
| P62081\|RS7_HUMAN | 4 | 4 | 0.84547 | 1.077 | R10W | WT | 40S ribosomal protein S7 GN=RPS7 |
| A7E2V4\|ZSWM8_HUMAN | 1 | 1 | 0.84552 | 1.125 | WT | R10W | Zinc finger SWIM domain-containing protein 8 GN=ZSWIM8 |
| P03243\|E1B55_ADE05 | 6 | 6 | 0.84590 | 1.052 | R10W | WT | E1B 55 kDa protein OS=Human adenovirus C serotype 5 OX=28285 |

| | | | | | | | |
|---|---|---|---|---|---|---|---|
| P30153\|2AAA_HUMAN | 2 | 2 | 0.84634 | 1.132 | WT | R10W | Serine/threonine-protein phosphatase 2A 65 kDa regulatory subunit A alpha isoform GN=PPP2R1A |
| P23396\|RS3_HUMAN | 6 | 6 | 0.84726 | 1.015 | WT | R10W | 40S ribosomal protein S3 GN=RPS3 |
| P63241\|IF5A1_HUMAN | 13 | 13 | 0.84728 | 1.029 | R10W | WT | Eukaryotic translation initiation factor 5A-1 GN=EIF5A |
| P06733\|ENOA_HUMAN | 21 | 15 | 0.85283 | 1.014 | WT | R10W | Alpha-enolase GN=ENO1 |
| Q9HCN8\|SDF2L_HUMAN | 1 | 1 | 0.85470 | 1.116 | WT | R10W | Stromal cell-derived factor 2-like protein 1 GN=SDF2L1 |
| P04264\|K2C1_HUMAN | 16 | 11 | 0.85477 | 2.367 | WT | R10W | Keratin, type II cytoskeletal 1 GN=KRT1 |
| P13639\|EF2_HUMAN | 30 | 29 | 0.85555 | 1.026 | R10W | WT | Elongation factor 2 GN=EEF2 |
| P25205\|MCM3_HUMAN | 1 | 1 | 0.85682 | 1.032 | WT | R10W | DNA replication licensing factor MCM3 GN=MCM3 |
| P18621\|RL17_HUMAN | 2 | 2 | 0.86048 | 1.019 | WT | R10W | 60S ribosomal protein L17 GN=RPL17 |
| P46777\|RL5_HUMAN | 3 | 3 | 0.86109 | 1.031 | R10W | WT | 60S ribosomal protein L5 GN=RPL5 |
| P01780\|HV307_HUMAN | 4 | 1 | 0.86439 | 1.113 | R10W | WT | Immunoglobulin heavy variable 3-7 GN=IGHV3-7 |
| P26641\|EF1G_HUMAN | 10 | 10 | 0.86630 | 1.020 | R10W | WT | Elongation factor 1-gamma GN=EEF1G |
| P60981\|DEST_HUMAN | 9 | 9 | 0.86867 | 1.005 | R10W | WT | Destrin GN=DSTN |
| Q9BSE5\|SPEB_HUMAN | 1 | 1 | 0.86882 | 1.042 | WT | R10W | Agmatinase, mitochondrial GN=AGMAT |
| P22626\|ROA2_HUMAN | 2 | 2 | 0.87549 | 1.008 | R10W | WT | Heterogeneous nuclear ribonucleoproteins A2/B1 GN=HNRNPA2B1 |
| P04075\|ALDOA_HUMAN | 1 | 1 | 0.88139 | 1.118 | R10W | WT | Fructose-bisphosphate aldolase A GN=ALDOA |
| Q9H857\|NT5D2_HUMAN | 2 | 2 | 0.88141 | 1.039 | WT | R10W | 5'-nucleotidase domain-containing protein 2 GN=NT5DC2 |
| P19338\|NUCL_HUMAN | 20 | 20 | 0.88237 | 1.021 | R10W | WT | Nucleolin GN=NCL |
| P30533\|AMRP_HUMAN | 2 | 2 | 0.88435 | 1.016 | R10W | WT | Alpha-2-macroglobulin receptor-associated protein GN=LRPAP1 |
| P14625\|ENPL_HUMAN | 3 | 2 | 0.88681 | 1.155 | R10W | WT | Endoplasmin GN=HSP90B1 |
| P16615\|AT2A2_HUMAN | 6 | 6 | 0.88776 | 1.145 | R10W | WT | Sarcoplasmic/endoplasmic reticulum calcium ATPase 2 GN=ATP2A2 |
| P39019\|RS19_HUMAN | 1 | 1 | 0.88948 | 1.046 | R10W | WT | 40S ribosomal protein S19 GN=RPS19 |
| P09382\|LEG1_HUMAN | 1 | 1 | 0.88998 | 1.114 | WT | R10W | Galectin-1 GN=LGALS1 |
| P27708\|PYR1_HUMAN | 8 | 8 | 0.89112 | 1.011 | R10W | WT | CAD protein GN=CAD |
| Q9BPW8\|NIPS1_HUMAN | 1 | 1 | 0.89324 | 1.028 | WT | R10W | Protein NipSnap homolog 1 GN=NIPSNAP1 |
| P02768\|ALBU_HUMAN | 6 | 5 | 0.89382 | 1.102 | R10W | WT | Albumin GN=ALB |
| P46776\|RL27A_HUMAN | 2 | 2 | 0.89699 | 1.070 | R10W | WT | 60S ribosomal protein L27a GN=RPL27A |
| Q15645\|PCH2_HUMAN | 1 | 1 | 0.89969 | 1.049 | R10W | WT | Pachytene checkpoint protein 2 homolog GN=TRIP13 |
| P98161\|PKD1_HUMAN | 1 | 1 | 0.90581 | 1.007 | WT | R10W | Polycystin-1 GN=PKD1 |
| Q92499\|DDX1_HUMAN | 1 | 1 | 0.90864 | 1.142 | WT | R10W | ATP-dependent RNA helicase DDX1 GN=DDX1 |
| O95881\|TXD12_HUMAN | 10 | 9 | 0.91021 | 1.013 | R10W | WT | Thioredoxin domain-containing protein 12 GN=TXNDC12 |
| P55209\|NP1L1_HUMAN | 4 | 4 | 0.91066 | 1.044 | WT | R10W | Nucleosome assembly protein 1-like 1 GN=NAP1L1 |
| P15880\|RS2_HUMAN | 4 | 4 | 0.91386 | 1.001 | R10W | WT | 40S ribosomal protein S2 GN=RPS2 |
| P61353\|RL27_HUMAN | 1 | 1 | 0.91486 | 1.014 | WT | R10W | 60S ribosomal protein L27 GN=RPL27 |
| Q9H3K6\|BOLA2_HUMAN | 5 | 5 | 0.92043 | 1.012 | R10W | WT | BolA-like protein 2 GN=BOLA2 |
| P62913\|RL11_HUMAN | 6 | 6 | 0.93048 | 1.019 | WT | R10W | 60S ribosomal protein L11 GN=RPL11 |
| Q9Y230\|RUVB2_HUMAN | 1 | 1 | 0.93591 | 1.296 | WT | R10W | RuvB-like 2 GN=RUVBL2 |
| P05091\|ALDH2_HUMAN | 1 | 1 | 0.94209 | 1.015 | R10W | WT | Aldehyde dehydrogenase, mitochondrial GN=ALDH2 |
| Q92945\|FUBP2_HUMAN | 4 | 4 | 0.94346 | 1.034 | R10W | WT | Far upstream element-binding protein 2 GN=KHSRP |
| Q96AG4\|LRC59_HUMAN | 5 | 5 | 0.94754 | 1.005 | WT | R10W | Leucine-rich repeat-containing protein 59 GN=LRRC59 |
| P34931\|HS71L_HUMAN | 56 | 1 | 0.94835 | 1.030 | WT | R10W | Heat shock 70 kDa protein 1-like GN=HSPA1L |
| Q9BQG0\|MBB1A_HUMAN | 2 | 2 | 0.94846 | 1.002 | WT | R10W | Myb-binding protein 1A GN=MYBBP1A |
| Q9Y679\|AUP1_HUMAN | 1 | 1 | 0.95135 | 1.030 | R10W | WT | Ancient ubiquitous protein 1 GN=AUP1 |
| P19474\|RO52_HUMAN | 8 | 8 | 0.95206 | 1.011 | WT | R10W | E3 ubiquitin-protein ligase TRIM21 GN=TRIM21 |
| P47914\|RL29_HUMAN | 3 | 3 | 0.95862 | 1.013 | R10W | WT | 60S ribosomal protein L29 GN=RPL29 |
| A0A075B6P5\|KV228_HUMAN;P01615\|KVD28_HUMAN | 35 | 3 | 0.96107 | 1.068 | WT | R10W | Immunoglobulin kappa variable 2-28 GN=IGKV2-28 |
| Q13263\|TIF1B_HUMAN | 1 | 1 | 0.96247 | 1.009 | WT | R10W | Transcription intermediary factor 1-beta GN=TRIM28 |
| Q7L592\|NDUF7_HUMAN | 1 | 1 | 0.96780 | 1.951 | R10W | WT | Protein arginine methyltransferase NDUFAF7, mitochondrial GN=NDUFAF7 |
| O14980\|XPO1_HUMAN | 1 | 1 | 0.96940 | 1.016 | R10W | WT | Exportin-1 GN=XPO1 |
| Q10567\|AP1B1_HUMAN | 86 | 27 | 0.96955 | 1.033 | R10W | WT | AP-1 complex subunit beta-1 GN=AP1B1 |
| Q9BU61\|NDUF3_HUMAN | 1 | 1 | 0.97376 | 1.028 | WT | R10W | NADH dehydrogenase [ubiquinone] 1 alpha subcomplex assembly factor 3 GN=NDUFAF3 |
| A0A0A0MRZ9\|LV552_HUMAN | 2 | 1 | 0.97483 | 1.011 | R10W | WT | Immunoglobulin lambda variable 5-52 GN=IGLV5-52 |
| P68431\|H31_HUMAN;Q16695\|H31T_HUMAN;Q71DI3\|H32_HUMAN | 4 | 4 | 0.97956 | 1.203 | WT | R10W | Histone H3.1 GN=H3C1 |
| Q9UBC2\|EP15R_HUMAN | 31 | 30 | 0.98048 | 1.102 | R10W | WT | Epidermal growth factor receptor substrate 15-like 1 GN=EPS15L1 |
| Q8TB37\|NUBPL_HUMAN | 1 | 1 | 0.98287 | 1.045 | WT | R10W | Iron-sulfur protein NUBPL GN=NUBPL |
| Q04837\|SSBP_HUMAN | 2 | 2 | 0.98487 | 1.052 | WT | R10W | Single-stranded DNA-binding protein, mitochondrial GN=SSBP1 |
| P55769\|NH2L1_HUMAN | 1 | 1 | 0.99063 | 1.033 | R10W | WT | NHP2-like protein 1 GN=SNU13 |
| P62826\|RAN_HUMAN | 4 | 4 | 0.99796 | 1.030 | WT | R10W | GTP-binding nuclear protein Ran GN=RAN |
| P14866\|HNRPL_HUMAN | 1 | 1 | 0.99953 | 1.045 | R10W | WT | Heterogeneous nuclear ribonucleoprotein L GN=HNRNPL |

**Supplementary Table 3. List of AP2$\sigma$2 interacting proteins.** Proteins shown in grey are significantly different ($p<0.05$) between the WT and R10W samples. NEC2 is a human herpesvirus 7 (strain JI) protein and was not included in the STRING analysis.
